# Burden is in the eye of the beholder: Sensitivity of yellow fever disease burden estimates to modeling assumptions

**DOI:** 10.1101/2021.01.06.21249311

**Authors:** T. Alex Perkins, John H. Huber, Quan Tran Minh, Rachel J. Oidtman, Magdalene K. Walters, Amir S. Siraj, Sean M. Moore

## Abstract

Geographically stratified estimates of disease burden play an important role in setting priorities for the management of different diseases and for targeting interventions against a single disease. Such estimates involve numerous assumptions, which uncertainty about is not always well accounted for. We developed a framework for estimating the burden of yellow fever in Africa and evaluated its sensitivity to assumptions about the interpretation of serological data and choice of regression model. We addressed the latter with an ensemble approach, and we found that the former resulted in a nearly twentyfold difference in burden estimates (range of central estimates: 8.4×10^4^-1.5×10^6^ deaths in 2021-2030). Even so, statistical uncertainty made even greater contributions to variance in burden estimates (87%). Combined with estimates that most infections go unreported (range of 95% credible intervals: 99.65-99.99%), our results suggest that yellow fever’s burden will remain highly uncertain without major improvements in surveillance.

## INTRODUCTION

Yellow fever is a mosquito-borne viral disease that poses a risk to people throughout tropical areas of South America and Africa [1]. The causative agent, yellow fever virus, is maintained in an enzootic cycle in non-human primates, and it infects humans primarily through spillover in communities in close proximity to sites of yellow fever epizootics in non-human primates [2]. Once infected, people experience a spectrum of disease severity, ranging from asymptomatic and mild infection to severe disease and death [3].

Thanks to safe and highly efficacious vaccines [4], yellow fever is vaccine-preventable in humans. Vaccinating the many people at risk of yellow fever on an ongoing basis is a challenge, however, given that areas where the virus occurs are geographically widespread and are inhabited by large populations with high birth rates [5]. The global supply of yellow fever vaccine is also a limiting factor, given that outbreak response contributes to the depletion of vaccine stockpiles above and beyond use of the vaccine for routine immunization and supplementary immunization activities [6].

In light of this complexity, modeling is an important tool for guiding vaccination policy for yellow fever. Models offer the ability to extrapolate beyond known reports of yellow fever to account for underreporting [3], to account for the influence of vaccination coverage, demographic structure, and natural immunity on incidence patterns [7,8], and to leverage spatial patterns in data to inform geographically realistic estimates [9]. After accounting for these factors to attain a model of disease burden, models can then be run under alternative scenarios about future vaccination to project its impact on the future burden of disease [9–12].

Several studies have modeled the probability of yellow fever occurrence (a binary outcome) [13–16], but only a few have explicitly modeled its burden (a continuous outcome) [9–12,17,18]. Collectively, these models span a range of assumptions, model structures, and inputs, any one of which can be viewed as reasonable and defensible. Uncertainty in these modeling choices has generally not been accounted for in burden estimates and impact projections, meaning that uncertainty therein may be underrepresented. Gaythorpe et al. have begun to address model uncertainty by taking weighted averages of models that represent alternative assumptions about transmission route [18] and spatial covariate data [12]. Numerous other forms of model uncertainty remain unexplored.

In this study, we explored how alternative assumptions about the interpretation of serological data and alternative approaches to regression modeling impact estimates of yellow fever burden in Africa and projections of future vaccination impact. Our modeling framework involves five sequential steps (Fig. 1) that first establish estimates of underreporting of cases and deaths informed by sites with serological data (Steps 1 & 2), next use estimates of underreporting to extrapolate reported cases and deaths to estimates of force of infection for all sites (Step 3), and then perform regression modeling of force of infection against spatial covariates to smooth over noise in extrapolations from reported cases and deaths (Steps 4 & 5). We considered eight different scenarios about the interpretation of serological data (Table 1) and eight different approaches to regression modeling, resulting in a total of 64 alternative models. For our final estimates, we distilled those 64 models down to a set of eight ensemble models: one for each scenario about the interpretation of serological data. Finally, we translated ensemble projections of force of infection into projections of deaths and deaths averted by vaccination.

**Table 1.** Eight scenarios about the interpretation of serological data. The eight scenarios (rows) were defined based on combinations of two alternative assumptions (shading) about each of four issues (columns). Gray shading indicates consistency with scenario 1.

| **Serology scenario** | **Vaccination status when reported as unvaccinated** | **Include studies not reporting vaccination status** | **Vaccination status when not reported** | **Include studies as part of an outbreak investigation** |
| --- | --- | --- | --- | --- |
| 1 | Unvaccinated | Yes | Vaccinated according to local coverage | Yes |
| 2 | Vaccinated according to local coverage | Yes | Vaccinated according to local coverage | Yes |
| 3 | Unvaccinated | No | NA | Yes |
| 4 | Unvaccinated | Yes | Unvaccinated | Yes |
| 5 | Unvaccinated | Yes | Vaccinated according to local coverage | No |
| 6 | Vaccinated according to local coverage | Yes | Vaccinated according to local coverage | No |
| 7 | Unvaccinated | No | NA | No |
| 8 | Unvaccinated | Yes | Unvaccinated | No |

**Figure 1.**
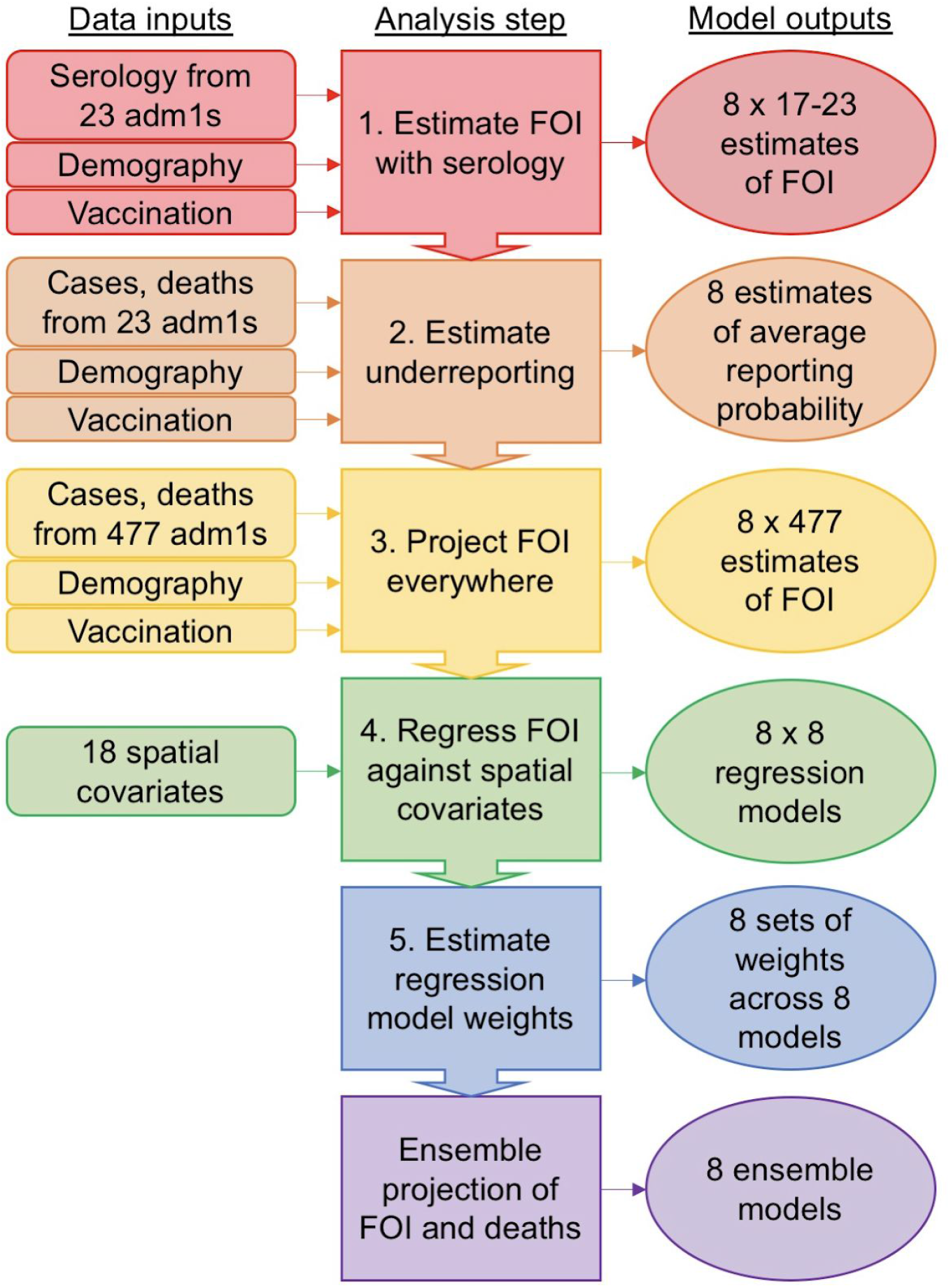
Modeling framework schematic. Our modeling framework involves five sequential steps that result in a set of eight ensemble models of the force of infection (FOI) of yellow fever virus, and associated deaths, in each of 477 first administrative-level units (adm1s) across 34 countries in Africa. Each of these eight ensemble models corresponds to a different assumption about the interpretation of serological data in Step 1 (Table 1). Colors associated with the six steps are used in subsequent figures to refer to the step to which those results pertain.

## RESULTS

### Step 1 – Estimate force of infection with serology

Estimates of force of infection (FOI) based on serology varied widely (Fig. 2) across the 23 sites for which serological data were available (Table S1) and across the eight assumptions about the interpretation of serological data that we considered (Table 1). Under serology scenario 1, estimates of FOI ranged from a median rate of 9.4×10^−7^ infections per susceptible person per year (95% CrI: 1.3×10^−8^-6.8×10^−4^) in Rift Valley Province, Kenya (22/433 seropositive across multiple age groups) to a median of 0.36 (95% CrI: 0.16-0.89) in Région du Nord, Cameroon (17/24 seropositive among 0-13 year olds). Posterior checks of predicted seropositives were consistent with the data on which the estimates were based (Fig. S1). Under scenarios 2 and 6, estimates of FOI at many sites were much lower (Fig. 2B, 2F), because those scenarios assumed that participants from all studies were vaccinated at levels consistent with age-specific coverage in that area in that year. That assumption resulted in much of the seropositivity being accounted for by prior vaccination, requiring a far lower FOI to explain the data. FOI estimates under scenarios 3 and 7 (Fig. 2C, 2G) were identical to scenario 1, except that some sites were dropped from the latter due to vaccination status of study participants being unknown or having been part of an outbreak investigation. Scenarios 4 and 8 (Fig. 2D, 2H) differed from scenarios 3 and 7 at only two sites, due to inclusion or exclusion of two studies in which the vaccination status of study participants was unknown.

**Figure 2.**
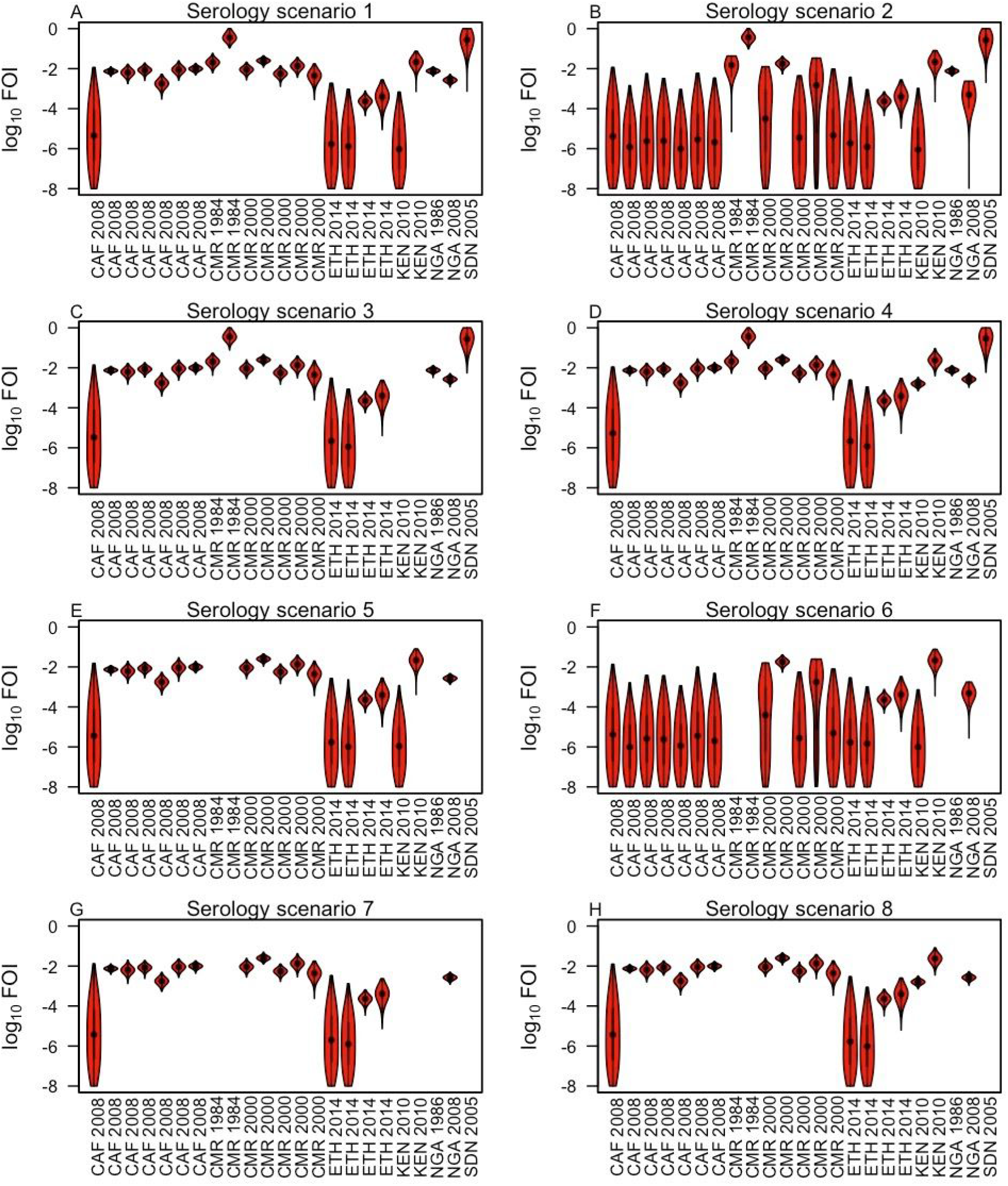
Estimated force of infection based on serology. The eight scenarios differ with respect to four assumptions about the interpretation of serological data (Table 1). Violin plots show the smoothed density of posterior samples obtained by Markov chain Monte Carlo. Country-year combinations are repeated along the x-axis for studies for which serological data was stratified sub-nationally. See Table S1 for more information about these studies.

### Step 2 – Estimate underreporting

Estimates of the probability that a yellow fever virus infection was reported varied several orders of magnitude across the 23 sites with serological data and the eight scenarios about the interpretation of serological data (Fig. 3). Rift Valley Province, Kenya had the highest estimated reporting probability (scenario 1: 95% credible interval: 0.031-0.044), and Kordofan, Sudan had the lowest (scenario 4: 95% CrI: 1.0×10^−8^-1.8×10^−7^) (Fig. 3A). In general, sites with large numbers of reported cases and deaths (large, filled circles in Fig. 3) had high estimates with narrow uncertainty, whereas those with zero reported cases and deaths (open circles in Fig. 3) had low estimates with wide uncertainty. To inform subsequent steps in our analysis, we calculated the average reporting probability across sites under each serology scenario. Scenarios 1, 2, 5, and 6 all had relatively high average reporting probabilities (highest = scenario 6: 2.5-3.5×10^−3^, 95% credible interval), and scenarios 3, 4, 7, and 8 all had relatively low average reporting probabilities (lowest = scenario 8: 1.3-2.3×10^−4^, 95% credible interval). This difference was due to either of two factors that differentiated these scenarios: inclusion of studies that did not report on the vaccination status of participants (scenarios 1 vs. 3, 5 vs. 7); or assumptions about the vaccination status of those study participants (scenarios 2 vs. 4, 6 vs. 8). Specifically, excluding studies that did not report on vaccination status of study participants or assuming that participants in those studies were not vaccinated led to higher estimates of force of infection (Fig. 2), which led to greater numbers of predicted infections and, consequently, lower estimates of reporting probability to explain the observed numbers of reported cases and deaths (Fig. 3).

**Figure 3.**
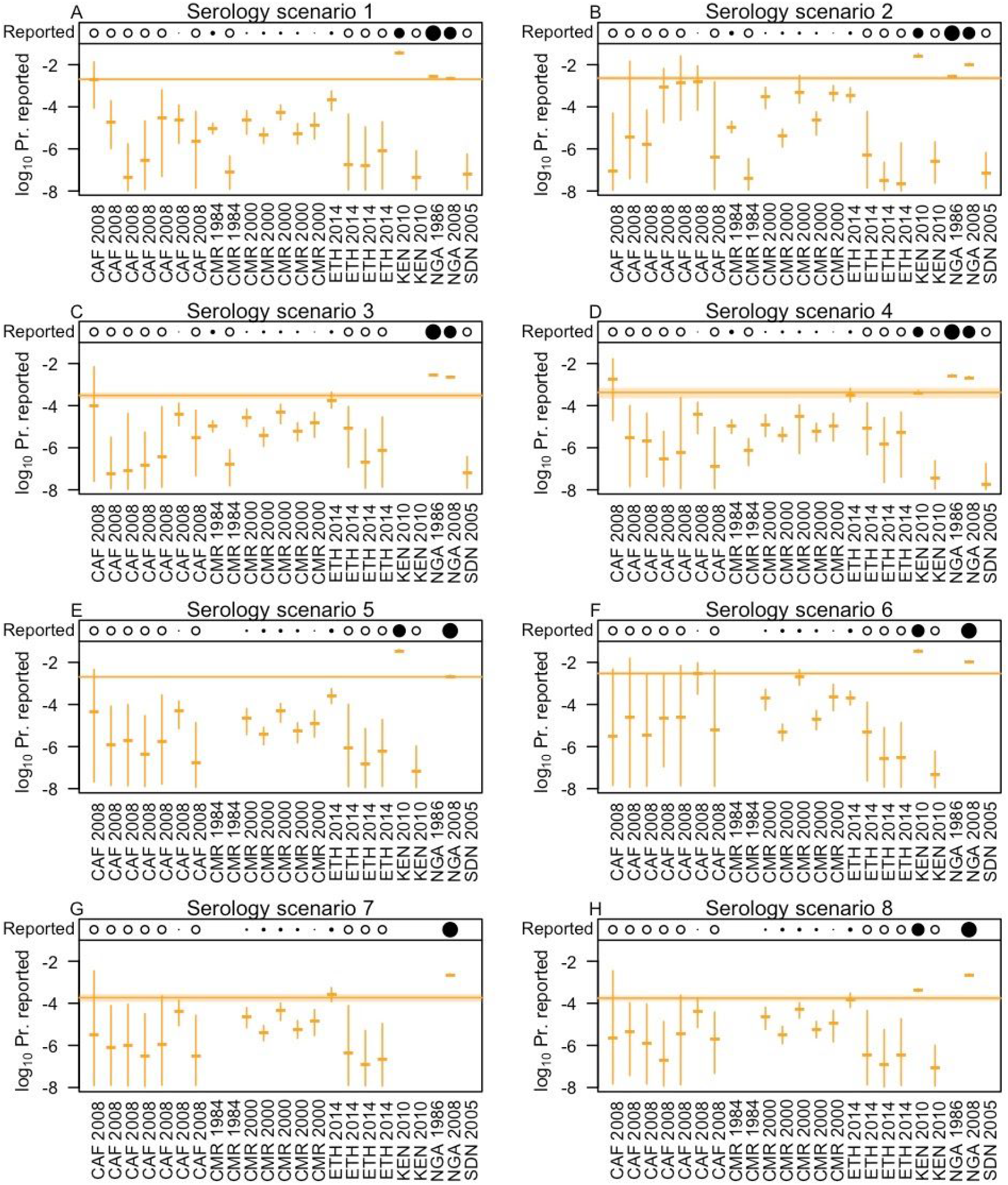
Estimated reporting probabilities. Dashes and line segments indicate median and 95% credible intervals of site-specific posterior estimates of the (log_10_) probability of a yellow fever virus infection being reported. Bands indicate posterior estimates of mean reporting probability across sites, averaged on a linear scale. The row along the top of each panel indicates the number of cases and deaths reported at each site during 1980-2014: open circle = zero; filled circle = log(reported cases + deaths); blank = not included. The eight scenarios in the panels and the 23 sites are the same as those in Fig. 2. See Table S1 for more information about these studies.

### Step 3 – Project force of infection everywhere

We estimated substantial spatial heterogeneity in FOI across the 477 first-level administrative units (adm1s) that we included in our analysis (Fig. 4A). Under serology scenario 1, estimates of force of infection (FOI) ranged from a low in Oromia, Ethiopia (95% CrI: 2.2×10^−8^-2.6×10^−6^) to a high in Grand Bassa County, Liberia (95% CrI: 0.25-3.9). The former has a large population (34 million), no vaccination coverage, and experienced zero cases or deaths during 1980-2014, whereas the latter has a small population (37,311), high vaccination coverage (91.8%), and experienced 360 cases and 9 deaths during that time frame (populations and vaccination coverages as of 2014, [8]). These were extremes though, with 90% of adm1s having a median FOI between 3.9×10^−6^ and 2.0×10^−3^. In general, adm1s with a low median FOI had high uncertainty (Fig. 4B), with their 95% credible intervals often spanning two orders of magnitude (Fig. 4C). In contrast, adm1s that reported positive numbers of cases or deaths during 1980-2014 (Fig. 4D) were associated with less uncertain estimates of FOI (Fig. 4B). This differential uncertainty was a consequence of the fact that there is an extremely wide range of values of FOI under which zero cases and deaths could be reported: i.e., low FOI with few infections, or high FOI with more infections but none reported. Overall, 64% of variance in FOI estimates was attributable to spatial heterogeneity, with the other 36% attributable to statistical uncertainty (see Supplemental Appendix for details of variance partitioning). Results from other serology scenarios had a similar spatial distribution but differed in magnitude consistent with differences in their estimated average reporting probabilities.

**Figure 4.**
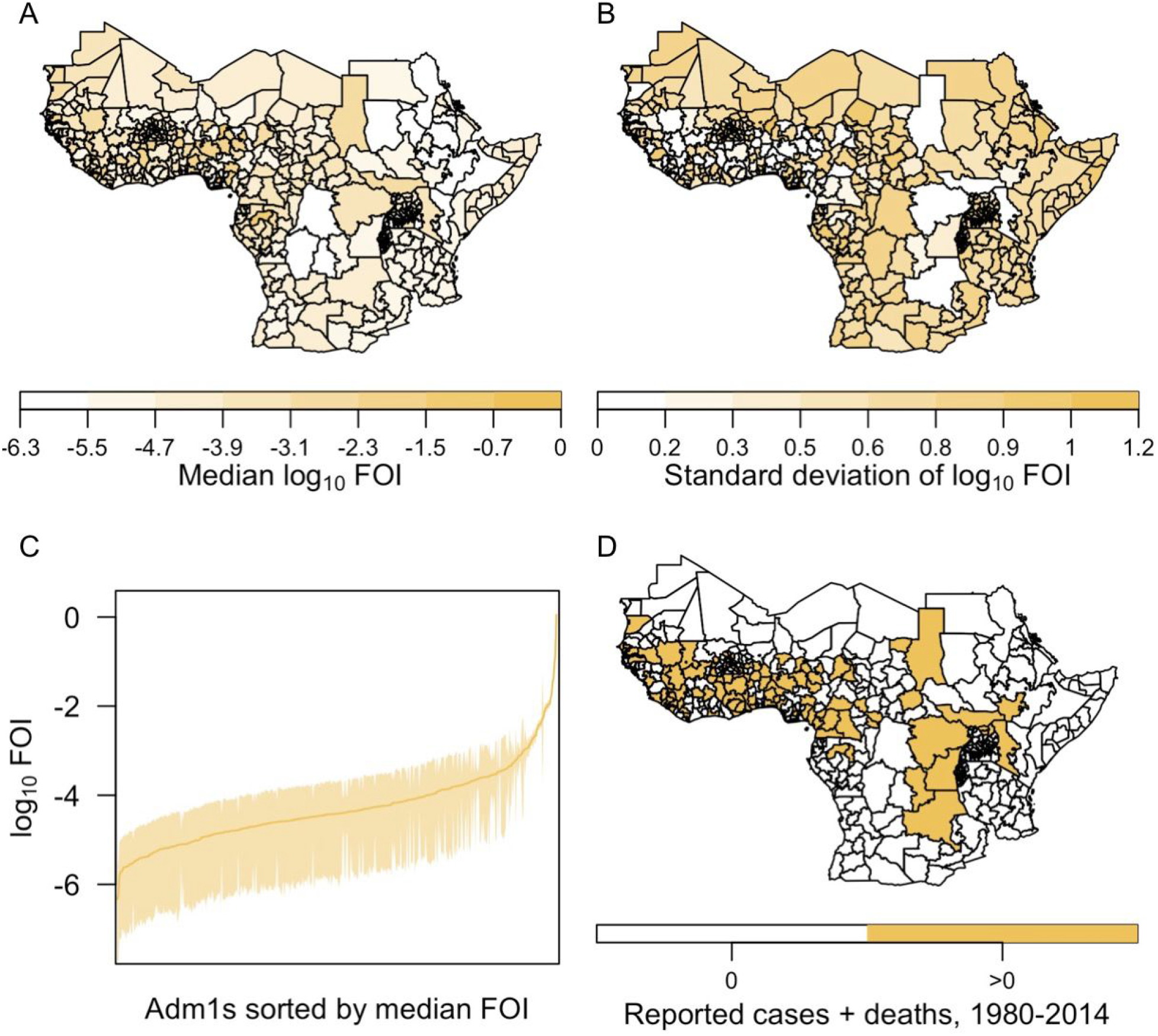
Force of infection projected from reported cases and deaths. Maps in A and B show the spatial distribution of the median and standard deviation of adm1-specific estimates of force of infection (FOI). The line and band in C show the median and 95% credible intervals of these estimates sorted by median FOI. Adm1s with reported cases or deaths in 1980-2014 are colored in D. Results presented here are in reference to serology scenario 1. Other serology scenarios had similar results, but with magnitude varying according to differences in the estimated average reporting probabilities shown in Fig. 3. Thus, results from serology scenarios 2, 5, and 6 were similar in magnitude to those presented here, whereas results from serology scenarios 3, 4, 7, and 8 were higher. All had a similar spatial distribution.

### Step 4 – Regress force of infection against spatial covariates

Regression of FOI projections from Step 3 against spatial covariates retained broad spatial patterns in FOI and brought extreme values of FOI toward the center of their range. In general, the regression models under serology scenario 1 resulted in median predictions of FOI across adm1s ranging 10^−6^-10^−3^ (Fig. 5), consistent with projections of median FOI from around 90% of adm1s from Step 3. These models accounted for 20-30% of variation in median values of projected FOI from Step 3 (Fig. S2). Markov random field models, which involve spatial smoothing, resulted in a somewhat narrower range of median FOI values, with the highest values across West and Central Africa (Fig. 5D-5F). Models that relied on more complex relationships among spatial covariates and FOI resulted in more spatially heterogeneous predictions across a wider range of median values of FOI (Fig. 5C, 5G). A linear model and boosted regression trees were more intermediate (Fig. 5B, 5H). As in Step 3, uncertainty around adm1-specific estimates of FOI was greater in adm1s with lower median values of FOI (Fig. S3). Results from other serology scenarios were similar but differed in magnitude consistent with differences in their estimated average reporting probabilities.

**Figure 5.**
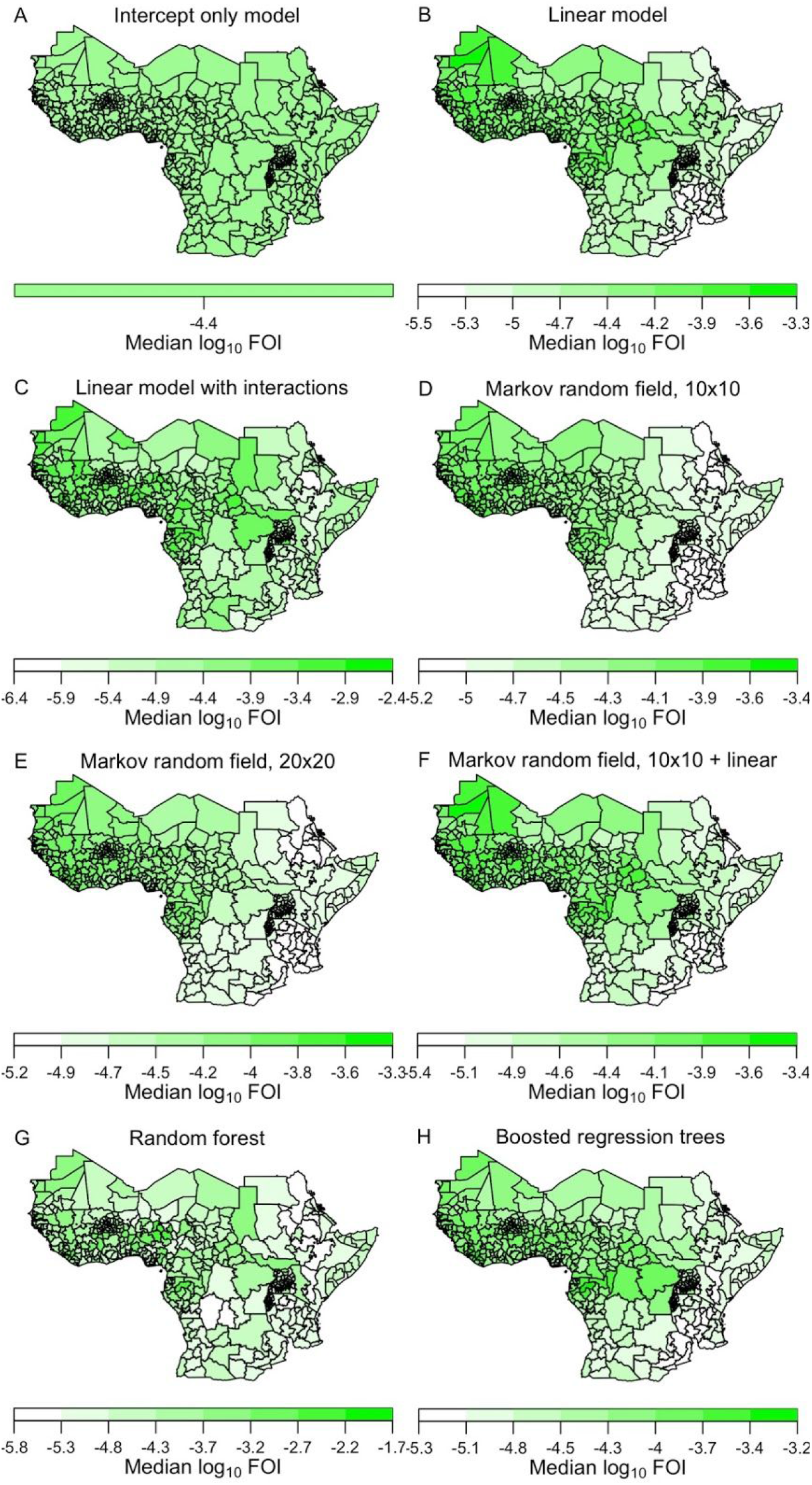
Spatial prediction of force of infection from eight regression models. Median values on a log_10_ scale are shown from serology scenario 1. Color axes for each model differ so as to maximize contrast within each panel. Other serology scenarios produced similar results, but with magnitude varying according to differences in the estimated average reporting probabilities shown in Fig. 3.

### Step 5 – Estimate regression model weights for ensemble model

Our process for generating an ensemble model involved assessing the performance of each regression model in 10-fold cross-validation, holding out three to four countries from model fitting and assessing predictions for first-level administrative units (adm1s) in those countries based on models fitted elsewhere (see Fig. S4 for the partition of countries we used for this). Relative to the intercept-only model (Int. in Fig. 6), much lower values of negative marginal log likelihood (NMLL) for the seven models that made use of spatial location and/or spatial covariates suggest that those variables have predictive value (Fig. 6A). On an individual basis, the linear model with interactions (Int.+) performed best under all serology scenarios (NMLL range: 991-1,575). This was primarily due to its high uncertainty relative to other models (Fig. S3), which allowed it to better cover the wide range of projected FOI values from Step 3. The ensemble model (Ens.) performed markedly better than all individual models (NMLL range: 605-673). On average across serology scenarios, the three Markov random field models (MRF10, MRF20, MRF10+) comprised 50% of the ensemble (range: 37-63%), boosted regression trees (BRT) 19% (range: 4-28%), the linear model (Lin.) 15% (range: 2-27%), and the intercept-only model 14% (range: 8-24%) (Fig. 6B). Despite performing well individually, the linear model with interactions and the random forest (RF) comprised very little of the ensemble, suggesting that their individual performances were bolstered by their high uncertainty rather than their ability to make accurate central predictions outside the data to which they were fitted. To enable the ensemble model to appropriately capture uncertainty in projected FOI from Step 3, it included an additional, Normally distributed noise term with a standard deviation of 0.79 (units: log_10_ FOI) on average across serology scenarios (range: 0.73-0.93).

**Figure 6.**
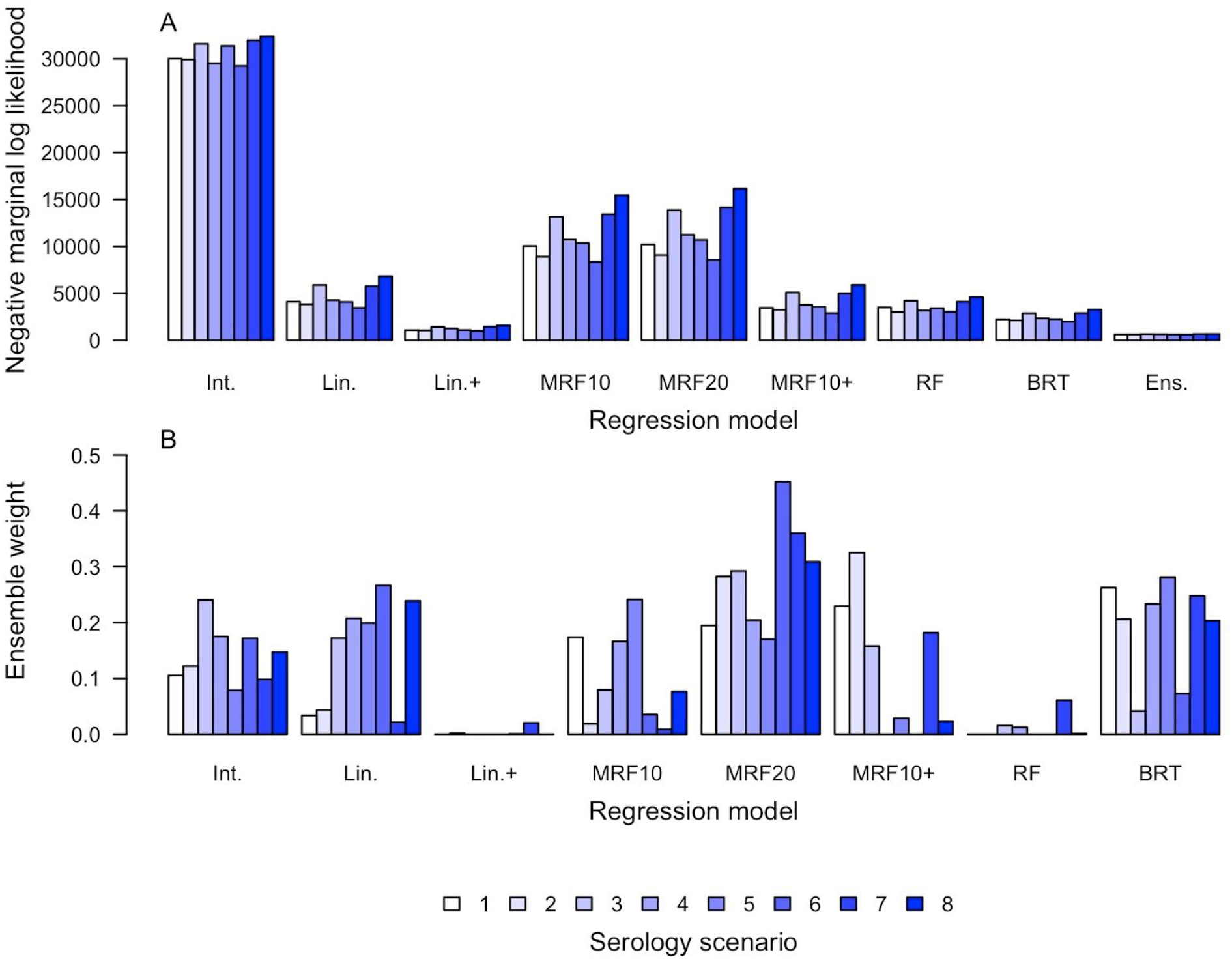
Regression model performance in cross-validation (top) and composition of the ensemble model (bottom). A) Performance in cross-validation was quantified using negative marginal log likelihood, with lower values indicating that the model was associated with a higher probability of generating the data withheld from fitting. For the purpose of this exercise, the “data” consisted of projected values of log_10_ force of infection from Step 3. Cross-validation was done in a 10-fold manner, with data from three to four countries withheld from fitting and used to assess out-of-fit prediction. B) Ensemble models consisted of linear combinations of adm1-specific predictions of log_10_ FOI, with the latter approximated by Normal distributions. The coefficients of those linear combinations (one for each serology scenario) are indicated by the height of the bars. In addition, the ensemble model included an additional Normal random variable with mean zero and standard deviation fitted as part of the process of constructing the ensemble.

### Ensemble projection of force of infection and deaths

The spatial distributions and ranges of median values of FOI under the ensemble models were broadly similar to projections arising from Step 3 (Fig. 7A). Specifically, median FOI was highest in West and Central Africa and ranged 5.1×10^−7^-3.0×10^−4^ under serology scenario 1. Uncertainty about FOI was generally lower in adm1s with high median values (Fig. 7B), although uncertainty was high across all adm1s due to the additional noise term in the ensemble model. On the log_10_ scale on which we modeled FOI, 64% of variance was attributable to statistical uncertainty, 25% to differences among serology scenarios, and 11% to spatial heterogeneity. On a linear scale, these proportions changed to 98.8%, 1.0%, and 0.2%, respectively. Under serology scenario 1, the spatial covariates that were most strongly associated with median values of log_10_ FOI were longitude (R^2^ = 0.69), one of the NDVI variables (R^2^ = 0.52), elevation (R^2^ = 0.44), one of the temperature variables (R^2^ = 0.36), latitude (R^2^ = 0.34), and two of the precipitation variables (R^2^ = 0.23-0.24) (Fig. S5). Due to collinearity among spatial variables, the apparent relationship between any given variable and FOI cannot be attributed entirely to that variable. Nonetheless, these associations provide an indication of the characteristics of adm1s associated with higher or lower forces of infection.

**Figure 7.**
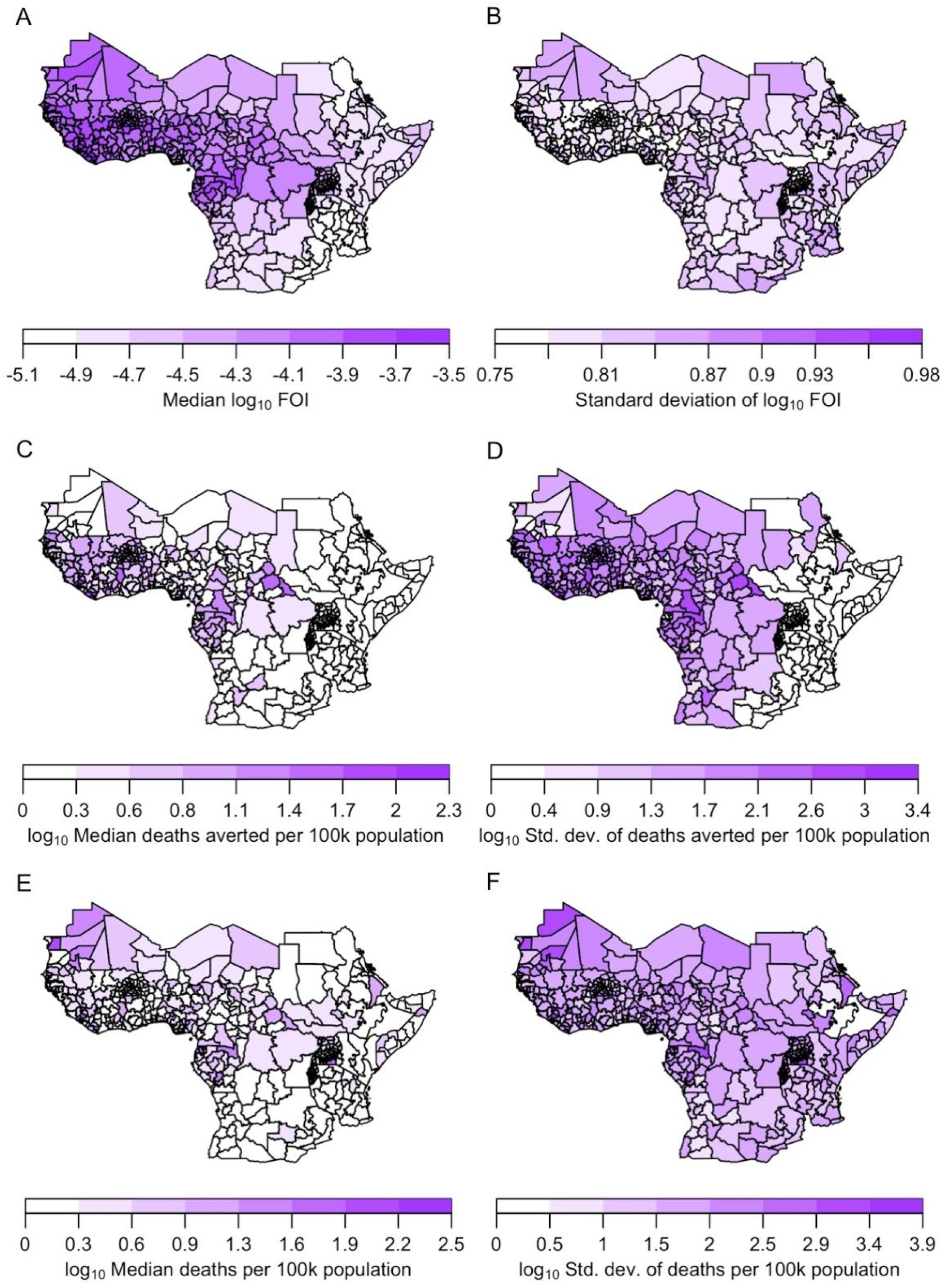
Ensemble model projections. Top: force of infection; middle: deaths averted by vaccination for 2021-2030; and bottom: deaths not averted. Left: median values; and right: standard deviation. All projections displayed here derived from serology scenario 1.

Projections of deaths averted during 2021-2030 were highest on a per population basis in adm1s in which FOI and vaccination coverage were both high (Figs. 7C, S6). Under serology scenario 1, vaccination was projected to avert deaths in 2021-2030 totaling 27,000 in Nigeria (95% posterior predictive interval: 3,500-135,000), 7,700 in Burkina Faso (95% PPI: 950-33,000), 6,900 in Côte d’Ivoire (95% PPI: 920-42,000), 5,000 in Ghana (95% PPI: 500-40,000), and 4,800 in Democratic Republic of Congo (95% PPI: 440-42,000) (Fig. S7). Under serology scenario 8, these projections were a full order of magnitude greater (Fig. S7). Despite that, only 8% of variance in projections of deaths averted was attributable to differences among serology scenarios, and only 7% to spatial heterogeneity across adm1s. The remaining 85% was attributable to statistical uncertainty. Relative to the earliest decade in our analysis (1980s), deaths averted by vaccination were projected to have increased by an order of magnitude (Fig. 8C) due to a combination of population growth (Fig. 8A) and increases in vaccination (Fig. 8B).

**Figure 8.**
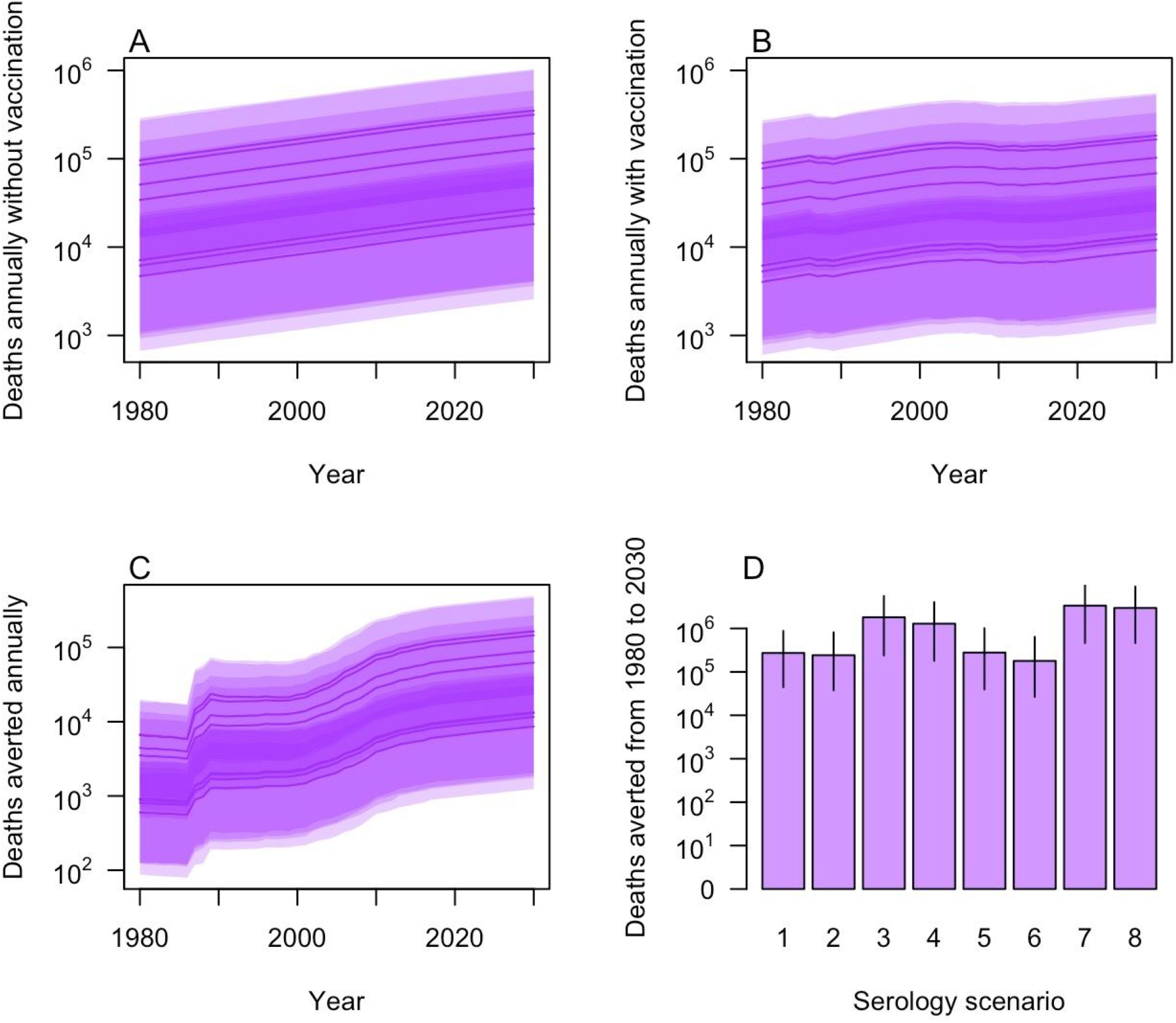
Deaths projected under the eight serology scenarios for 1980-2030. Deaths are presented: A) annually without vaccination; B) annually with vaccination; C) annually as the number averted by vaccination; and D) cumulatively. In A-C, projections under each serology scenario are presented as lines (median) and bands (95% posterior predictive interval).

Despite the large number of deaths averted by vaccination projected for 2021-2030, we projected a 74% chance that the number of deaths not averted will exceed the number that are averted in 2021-2030. The number of deaths in 2021-2030 not averted by vaccination ranged from 84,000 (95% CrI: 12,000-290,000) under serology scenario 6 to 1.5 million (95% CrI: 230,000-4.8 million) under serology scenario 8. On a per population basis, deaths in 2021-2030 were projected to occur both in adm1s with high deaths averted and in additional adm1s on the periphery of the most heavily vaccinated areas (Fig. 7E). Only 6% of variance in projected deaths in 2021-2030 was attributable to spatial heterogeneity, with 7% due to differences among serology scenarios and 87% attributable to statistical uncertainty.

## DISCUSSION

We developed a framework for making geographically stratified estimates of the burden of yellow fever in Africa and used it to assess the sensitivity of our estimates to two key model uncertainties. Our framework has similarities to another introduced by Garske et al. [9] (and expanded on in several ways since [10–12,18]), due in large part to the nature of data available to estimate yellow fever’s burden (i.e., serological data from a few locations, outbreak data from many). One distinction of our approach is that it makes use of information on the magnitude of reported cases and deaths, whereas the one by Garske et al. [9] makes use of the occurrence of reported cases or deaths only. As a result, our framework makes use of additional information not leveraged by Garske et al. [9]. At the same time, the accuracy of those data are questionable given significant challenges with yellow fever surveillance [19,20], although we account for underreporting and uncertainty therein in our approach. On average, differences arising from this choice may be limited. Under serology scenario 1, we projected 9,500 (95% posterior predictive interval: 1,600-31,000) deaths averted across Africa in 2018, as compared to 10,000 (95% credible interval: 6,000-17,000) in the most recent estimates by Gaythorpe et al. [12]. The most notable difference between these projections may be, therefore, in terms of their uncertainty rather than their central tendency, at least under similar assumptions.

A central focus of our analysis was the sensitivity of burden estimates to different assumptions about the interpretation of serological data. We found that the most consequential assumption was in regards to the vaccination status of participants in serological studies. A majority of serological studies that we included (21/23) reported that study participants had not been vaccinated. In the event that vaccination status was misreported or not recalled correctly [21–23], force of infection could have been much lower than if study participants had not been vaccinated, given that vaccination and natural infection are both capable of generating a positive serological result [24]. We found that this issue, even if it affects only a small number of serological studies, can be highly consequential for estimates of average reporting probability. This sensitivity propagated throughout the steps of our analysis, resulting in an order of magnitude difference in our central estimates of burden. Even so, the effect of this assumption on uncertainty was relatively minor (6% of variance in deaths projected for 2021-2030) compared to the much greater influence of statistical uncertainty (87%). The spatial distribution of burden was also generally similar across serology scenarios. As such, different assumptions about the interpretation of serological data may be more consequential for decision making around the prioritization of investments across different vaccine-preventable diseases [25] than for decision making that is limited in scope to yellow fever.

The other major focus of our analysis was the sensitivity of burden estimates to different approaches to regression modeling. We found that, individually, different regression models produced results in ways that were mostly predictable based on the usual tendencies of those methods. For example, Markov random field models produced median estimates that were smoother across space and associated with less uncertainty, as compared to models that allowed for complex effects of a set of 18 spatial covariates. We found that, collectively, an ensemble model composed of predictions from Markov random field models, linear models, and boosted regression trees performed best in cross-validation. Ours are the first burden estimates for yellow fever that take into account structurally distinct regression models and do so based on performance in cross-validation, although Gaythorpe et al. have recently developed ensembles composed of models that differ with respect to assumptions about transmission route [18] and sets of spatial covariates [12]. Our work adds to a growing set of studies that demonstrate the benefits of ensemble modeling for geographically stratified estimates of disease burden [26–30], in addition to other applications in infectious disease epidemiology [31–34].

As important as the aforementioned assumptions were, their contribution to overall uncertainty in our estimates was relatively small compared to that of statistical uncertainty. In Step 3 of our analysis, considerable variance in log_10_ force of infection was attributable to spatial heterogeneity (29%), serology scenario (51%), and regression model choice (12%). Two things diminished the amount of variance attributable to those factors in later steps in our analysis. First, the ensemble models required an additional noise term to perform well in cross-validation, which increased the proportion of variance in log_10_ force of infection attributable to statistical uncertainty from 8% to 64%. Second, transforming log_10_ force of infection to a linear scale disproportionately increased variance associated with statistical uncertainty, from 64% to 98.8% of total variance. Translation of force of infection into deaths then diminished the contribution of statistical uncertainty somewhat, likely due to the saturating relationship between force of infection and deaths. Even so, 85% of variance in deaths averted and 87% of variance in deaths was attributable to statistical uncertainty, which calls into question the extent to which geographic differences in yellow fever’s burden are predictable in the first place.

Given our estimates that only one in a thousand to one in ten thousand infections were reported, there may be limits to the extent that improved modeling can reduce uncertainties about yellow fever’s burden. Increasing surveillance and diagnostic capacity—which is already an emphasis of efforts to prevent yellow fever epidemics [35]—could help reduce underreporting and, thereby, uncertainty in burden estimates. Our method is well-suited to leverage any such future improvements in surveillance data, given that it makes use of information about numbers of cases and deaths as opposed to occurrence only. Additional serological data could also help resolve uncertainties in burden estimates. Most critically, if surveys could be conducted with serological assays that are capable of distinguishing between vaccine-derived and naturally acquired immunity (which is not currently the case [24]), that would greatly reduce uncertainty associated with the eight serology scenarios we considered. Even in the absence of assays with that capability, additional serological surveys would strengthen confidence in our ability to extrapolate reporting probability estimates from Step 2 to region-wide projections in Step 3.

Doing so in a manner that randomizes site selection and standardizes criteria for recruitment of study participants would be ideal.

Our analysis addressed the extent to which two types of model assumptions influence burden estimates, but there are others we did not explore that could be important. First, in addition to infections resulting from zoonotic spillover, there is also a role of urban transmission of yellow fever virus in Africa that we did not account for [10,36,37]. In a recent comparison of alternative models premised on zoonotic spillover versus urban transmission, the former was found to much better explain available data from Africa [18]. Second, because of the episodic nature of urban outbreaks in humans and epizootics in non-human primates [2], models that account for inter-annual variability in force of infection (e.g., [38]) could better match the realities of yellow fever’s epidemiology. To date, other models of yellow fever’s burden have not addressed this issue either [9–12,17,18], and doing so could be challenging given the widespread perception that reporting of yellow fever outbreaks is extremely sparse [9,39]. Third, we assumed a fixed value of lifelong protection from vaccination, which has recently been called into question based on waning antibody titers in vaccine recipients over time [40,41]. This could be an issue to investigate further in future work, but long-lasting protection remains consistent with prevailing assumptions about yellow fever vaccines [42]. There are also uncertainties about vaccination coverage and demography that we did not address but that could be important [7,17].

## CONCLUSION

Although we did not consider every possible model variant imaginable, our analysis made an important advance in demonstrating how alternative modeling assumptions can be accounted for in burden estimates for yellow fever. In the future, models that make the same assumptions about the interpretation of data but differ in their assumptions about drivers of transmission could be accommodated under our approach to ensemble modeling. When models make different assumptions about the interpretation of data, they may not be combinable under our ensemble approach (if their likelihoods are not comparable), but their contribution to overall uncertainty can be quantified nevertheless, as we demonstrated. Doing so illustrated that, while advances in modeling are important, improvements in data quality will be necessary to improve estimates of yellow fever’s burden and projections of the impacts of vaccination thereon.

## METHODS

### Data

Spatially, our analysis focused on 34 countries in Africa considered endemic or at risk for yellow fever, and for which necessary demographic and vaccination coverage estimates were available [8]. Temporally, we focused on the period from 1980 to 2014. We made this determination based on the last year of data available in one of the epidemiological datasets we used.

All phases of our analysis used estimates of population and vaccination coverage by Hamlet et al. [8] that were stratified by age, year, and first subnational administrative unit (adm1). National totals for population by age derived from United Nations World Population Prospects estimates [43], and spatial disaggregation thereof among adm1s was done based on LandScan 2015 estimates [44]. Vaccination coverage estimates compiled information from a variety of sources dating back as long ago as the 1940s on routine immunization, reactive vaccination, and preventive mass immunization campaigns [8]. These estimates can be perused in full detail at https://shiny.dide.imperial.ac.uk/polici/. Average values of population and vaccination coverage during 2021-2030 for each adm1 are displayed in Fig. S6.

The regression phase of our analysis made use of data on several spatial variables thought to be associated with yellow fever, each of which was based on raster data that we averaged across the first administrative level. These variables include normalized difference vegetation index (NDVI) [45], monthly precipitation [46], monthly average temperature [46], elevation [47], longitude, latitude, travel time to the nearest urban center [48], occurrence probability [17] and richness [49] of non-human primate (NHP) species known to be yellow fever virus reservoirs in Africa, percentage of frontier and tropical land cover [50], and forest loss [51]. We also used the health access quality index [52] at the national level. To reduce the dimensionality of monthly data, we performed principal components analyses on monthly NDVI, precipitation, and temperature data using the prcomp function in R [53]. We retained principal components explaining >95% of variation in total, resulting in two principal components for NDVI, four for precipitation, and two for temperature (see Fig. S8 for loadings). Prior to regression modeling, we centered and scaled all predictor variables (see Fig. S9 for maps of centered and scaled values of each variable).

We used two types of epidemiological data in our analysis. The first was derived from 23 published serological surveys [54–61], which captured information about past exposure to yellow fever virus. We recorded the number tested and number positive for each age group reported, and we limited our analysis to neutralization assays to maximize specificity of test results. Attributes of the sites where these serological surveys were conducted are detailed in Table S1. Second, we used data on yellow fever outbreaks compiled and shared with us by Garske et al. from [9]. This included data on cumulative cases and deaths that were reported at the adm1 level over the period of our analysis. The sources of these data were the WHO Weekly Epidemiological Record [62] and WHO Disease Outbreak News [63]. These data spanned 236 unique adm1-year combinations and included a total of 19,550 reported cases (of which 427 were confirmed) and 4,887 reported deaths (of which 37 were confirmed), which is fewer than the 32,731 cases reported at a national level by WHO during the study period [64]. Without an empirical resolution to this discrepancy, we relied on serological data to combine with these data to inform estimates of a reporting probability that accounts for this and other forms of underreporting. For confirmed cases and deaths, the method of confirmation was indicated as an IgM enzyme-linked immunosorbent assay in most instances, sometimes in combination with either a reverse transcription polymerase chain reaction test or a plaque reduction neutralization test. For reported cases and deaths, information about clinical diagnostic criteria were not specified [62,63].

### Framework for estimating disease burden

#### Step 1 - Estimate force of infection with serology

For each administrative unit where serological data were available, we obtained a probabilistic estimate of a temporally constant force of infection (FOI) for that site. For a survey conducted in administrative unit *i* in year *y*, we calculated the likelihood of the FOI at *i, FOI*_*i*_, based on the number of individuals between ages *a*_1_ and *a*_2_ who tested positive, 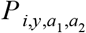, among the number who were tested, 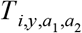. We assumed that 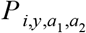 ∼ Binomial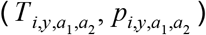, where 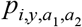 is the probability that an individual was seropositive. Our formulation accounted for the possibility that individuals could be seropositive due either to prior exposure to yellow fever virus or due to vaccination. To account for this, the probability of being seropositive was defined as

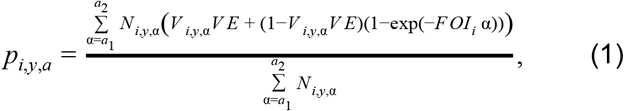

where *V*_*i,y,a*_ is vaccination coverage, *N*_*i,y,a*_ is population, and *VE* is vaccine efficacy, which we assumed to be 0.975 [4]. We used estimates of *V*_*i,y,a*_ and *N*_*i,y,a*_ produced by Hamlet et al. [8]. We calculated the log likelihood of each *FOI*_*i*_ by summing the logs of the binomial probabilities of 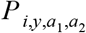 across all age groups. Using this log likelihood and a uniform prior between 10^−8^ and 1, we sampled from the posterior distribution of each *FOI*_*i*_ using the BayesianTools [65] package in R [53]. We used the default DEzs sampler, running three chains for a total of 10^4^ iterations and applying a burnin of 10^3^ iterations. We assessed convergence by calculating the multivariate potential scale reduction factor and verifying that it was near one.

#### Step 2 - Estimate underreporting

For each administrative unit where serological data were available, we estimated the extent of underreporting based on the discrepancy between observed cases and deaths and the number of infections predicted by the FOI estimates from those sites. This analysis was centered around the distribution of person-years across three categories: observed deaths, *D*; observed cases, *C*; and unobserved person-years among individuals unprotected by vaccination, *N*.

For administrative unit *i*, the probability that a person of age *a* in year *y* who was unprotected by vaccination would die from yellow fever and be reported as such was

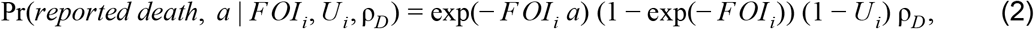

where *U*_*i*_ is the proportion of infections that are unobserved in *i* and ρ_*D*_ is the proportion of observed infections that result in death. The probability of a reported case was the same as eqn. 2 but with ρ_*D*_ replaced by 1 − ρ_*D*_. The probability of an unobserved person-year allowed for multiple ways in which a person-year would not result in a reported death or case, including by having been infected previously, by never being infected, or by being infected but not being reported. This resulted in

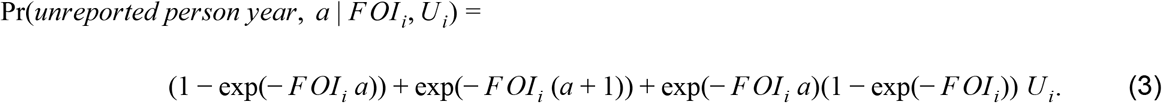

With these three probabilities, we were able to calculate the probability of *D*_*i*_, *C*_*i*_, and *N*_*i*_ among *Y*_*i*_ total person-years, Pr(*D*_*i*_, *C*_*i*_, *N*_*i*_ | *FOI*_*i,j*_, *U*_*i*_, ρ_*D*_), using a multinomial distribution with component probabilities defined by eqn. 2, substitution of *D* with *C* in eqn. 2, and eqn. 3. The quantity *Y*_*i*_ included all individuals across ages *a* in all applicable years who had not been vaccinated or, with probability 1-*VE*, those who had been vaccinated. This was used to define *N*_*i*_ as *Y*_*i*_ - *C*_*i*_ - *D*_*i*_.

The parameters we sought to estimate based on these data were *U*_*i*_ for each *i* and a single value of ρ_*D*_ that was common to all *i*. To incorporate the full posterior distribution of each *FOI*_*i*_ estimated in Step 1, we calculated the marginal probability of the data,

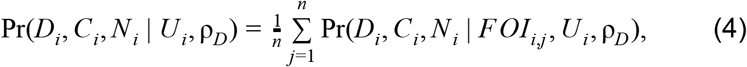

by averaging over uncertainty in *FOI* from its posterior distribution, the *n* = 10^3^ samples of which are indexed by *j*. We calculated the log likelihood of *U*_*i*_ and ρ_*D*_ by summing the logs of the probabilities from eqn. 4 across all *i*. We assumed noninformative priors between 0 and 1 for all *U*_*i*_ and a beta-distributed prior for ρ_*D*_ with shape parameters 2.05 and 6.85, which were informed by previous estimates [3] (see Supplemental Appendix). We sampled from the posterior distributions of the parameters using the BayesianTools [65] package in R [53]. We used the default DEzs sampler, running three chains for a total of 3×10^4^ iterations, applying a burnin of 5×10^3^ iterations, and thinning to retain every fifth iteration. We assessed convergence by calculating the multivariate potential scale reduction factor and verifying that it was near one.

To allow for extrapolation of underreporting beyond the few administrative units with serological data, we fitted a Dirichlet distribution to posterior predictions of the proportions of infections that result in a reported death, a reported case, or an unreported infection. In doing so, we took an average across sites *i* for each draw *j* from the posterior, such that 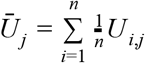. For each draw *j* from the posterior, these proportions were calculated as (1 − *Ūj*) ρ_*D,j*_, (1 − *Ūj*) (1 − ρ_*D,j*_), and *Ūj*, and the Dirichlet parameters associated with them were α_*D*_, α_*C*_, and α_*U*_. We estimated these Dirichlet parameters by maximum likelihood using the optim function in R [53], treating posterior predictions of the proportion of infections that result in a reported death, a reported case, or an unreported infection as data points drawn from the Dirichlet distribution being fitted. In that sense, the Dirichlet distribution was a parametric approximation of the posterior samples of (1 − *Ūj*) ρ_*D,j*_, (1 − *Ūj*) (1 − ρ_*D,j*_), and *Ūj*, which was more convenient to work with than posterior samples in the next step of our analysis.

#### Step 3 - Project force of infection everywhere

For all administrative units, we estimated the total number of infections that occurred, *I*_*i*_, based on numbers of reported deaths and reported cases, together with the proportions of reported deaths, reported cases, and unobserved infections estimated in Step 2. The first step in this process was to calculate the maximum number of infections, *I*_*i,max*_, that could have possibly occurred in *i*. To do so, we calculated the number of infections that would have occurred if *FOI*_*i,max*_ = 10, which is well above what we considered to be a plausible value and above which *I*_*i,max*_ would not have been measurably larger. Across all ages *a* and years *y*, the total number of infections that would have resulted from *FOI*_*i,max*_ was

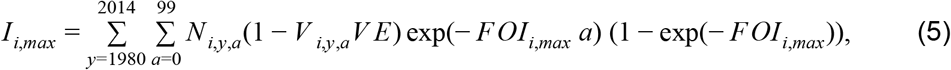

where *N*_*i,y,a*_ is population. We considered the minimum number of infections, *I*_*i,min*_, to be the sum of reported deaths, *D*_*i*_, and reported cases, *C*_*i*_. Based on this, we calculated the likelihood of a given number of infections,

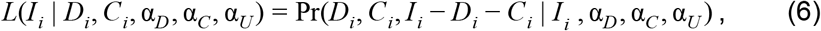

as the Dirichlet-multinomial probability of obtaining *D*_*i*_ reported deaths, *C*_*i*_ reported cases, and *I*_*i*_ - *D*_*i*_ - *C*_*i*_ unobserved infections following *I*_*i*_ draws from those categories according to Dirichlet-distributed probabilities with parameters α_*D*_, α_*C*_, and α_*U*_ from Step 2. We normalized the likelihoods from eqn. 6 across all values of *I*_*i*_ to obtain posterior probabilities of each *I*_*i*_, which we used to obtain a set of posterior samples of *I*_*i*_ by sampling with replacement from *I*_*i,min*_ to *I*_*i,max*_ proportional to the posterior probability of each *I*_*i*_.

For all administrative units, we translated estimates of the total number of infections, *I*_*i*_, into estimates of force of infection, *FOI*_*i*_. For each sample of *I*_*i*_ from Step 3, we found the value of *FOI*_*i*_ that minimized the absolute value of the difference between *I*_*i*_ and the expected number of infections under *FOI*_*i*_, which was

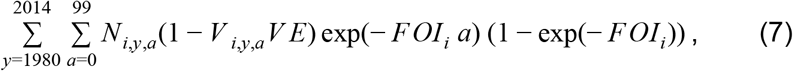

using the optimize function in R [53]. This resulted in a set of posterior samples of *FOI*_*i*_ for each administrative unit.

#### Step 4 - Regress force of infection against spatial covariates

Although Step 3 provides probabilistic estimates of force of infection that could be used to quantify disease burden, these estimates were highly sensitive to numbers of reported deaths and cases, which are noisy signals of underlying transmission. To smooth across that noise and obtain a more robust description of spatial patterns of FOI, we performed regression analyses of log_10_ FOI against a set of spatial variables. For each of eight regression models (the details of which are described later in the Methods), we performed separate regressions on each of 10^3^ samples of log10 *FOI i* from all administrative units. This resulted in 10^3^ replicate regressions and a set of 10^3^ predicted values of log10 *FOI i* from each regression model. We used the point estimates from each regression model as predicted values, allowing uncertainty to be accounted for through variability across replicate regressions. This is consistent with the property of equivariance, which allows quantities derived from posterior samples of parameters to themselves be considered posterior samples [66]. Thus, the set of predicted values of log_10_ *FOI*_*i*_ for site *i* from a given regression model constituted a posterior set of point estimates associated with that regression model.

#### Step 5 - Estimate regression model weights for ensemble model

For each serology scenario, we generated an ensemble model projection of FOI in each adm1 using a form of stacked generalization [26]. This approach regards the eight regression models as being at one level and seeks to generate another model at a higher level that weights the predictions of the eight models into its own prediction. A model at this higher level is considered successful if its set of predictions, 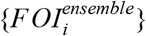, match the set of estimates from Step 3, 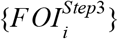, in cross-validation. The starting point for these predictions are predictions of data withheld from fitting of each model *m*, which we denote 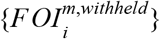. On a log scale, these predictions were approximated reasonably well by a normal distribution with parameters μ_*i,m*_ and σ_*i,m*_, which we estimated based on maximum-likelihood. We defined our ensemble predictions by another Normal distribution that represents a weighted average of the separate model predictions and has parameters μ_*i,ensemble*_ = Σ_*m*_α_*m*_μ_*i,m*_ and 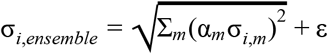, where Σ_*m*_α_*m*_ = 1, all α_*m*_ > 0, and ε >0. We informed estimates of the model weights, {α_*m*_}, on the basis of the marginal likelihood,

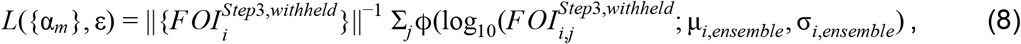

where ϕ denotes the Normal probability density function and *j* is an index of replicates over which the likelihood is marginalized. As indicated in the superscript, 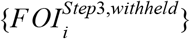 represents estimates of FOI from Step 3 that were withheld from model fitting. We partitioned data first for model fitting and then for ensemble model fitting at the country level, with ten different partitions of countries in which each partition used approximately 90% of administrative units for model fitting and approximately 10% of administrative units for cross-validation (Fig. S4). This partitioning was determined so as to maximize the evenness of the number of adm1s across partitions. To do this, the first partition took the country with the most adm1s and the two countries with the fewest and grouped them together. The second partition took the country with the second most adm1s and the two countries with the third and fourth fewest. This process was repeated until ten partitions of three to four countries each were obtained. To estimate {α_*m*_} and ε, we performed a constrained optimization to identify values of {α_*m*_} and ε that minimized a negative marginal log likelihood (NMLL) based on eqn. 8 using the constrOptim function in R [53], subject to the constraints on α_*m*_ and ε stated above.

#### Ensemble projection of force of infection and deaths

Using regression model weights, {α_*m*_}, and the additional noise term, ε, for the ensemble model, we drew 10^3^ Monte Carlo samples of 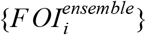 from Normal distributions with means μ*i,ensemble* and standard deviations σ_*i,ensemble*_. For each value *FOI*_*i,j*_ from 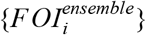, we computed the expected number of infections at each site *i* in year *y* as

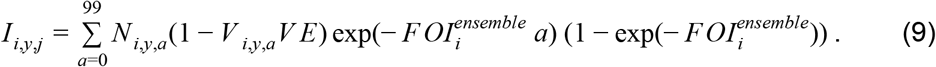

To calculate associated deaths, *D*_*i,y,j*_, we multiplied each *I*_*i,y,j*_ by a draw from a beta distribution with shape parameters 2.05 and 15.5, which were informed by previous estimates [3] (see Supplemental Appendix). This probability of death differed from ρ_*D*_ from Steps 2 and 3, because it pertained to all infections rather than just symptomatic infections. Deaths averted were calculated by taking differences between values of *D*_*i,y,j*_ calculated under different scenarios about *V*_*i,y,a*_.

### Alternative models

#### Interpretation of serological data

There were two general aspects of the interpretation of serological data to which we evaluated the sensitivity of our results. Alternative choices about these assumptions resulted in a total of eight distinct scenarios summarized in Table 1.

The first aspect of the interpretation of serological data that we considered was the vaccination status of study participants. Some studies were described as being performed on individuals with no prior vaccination against yellow fever, whereas descriptions of other studies did not specify this. Given that recall of vaccination status can be subject to considerable error [21–23], we assessed the sensitivity of our results to uncertainty that this leaves about the true vaccination status of study participants. Specifically, we considered four possibilities: 1) believe studies claiming that participants were unvaccinated and assume that participants in other studies were vaccinated consistent with local coverage (scenarios 1 & 5); 2) assume that participants from all studies were vaccinated consistent with local coverage (scenarios 2 & 6); 3) believe studies claiming that participants were unvaccinated and exclude other studies (scenarios 3 & 7); or 4) assume that participants from all studies were unvaccinated (scenarios 4 & 8). Under scenarios in which we assumed that participants were unvaccinated, this amounted to setting *V*_*i,y,a*_ = 0 in eqn. 1.

The second aspect of the interpretation of serological data that we considered was whether a survey was conducted as part of an outbreak investigation. Such surveys were omitted in the analysis by Garske et al. [9] due to concern that they would not be representative of force of infection at locations where no outbreak investigation had occurred. At the same time, there are a very limited number of serological surveys available, making any survey potentially valuable. To assess the sensitivity of our results to inclusion of these surveys, we crossed the four aforementioned scenarios about the vaccination status of serological survey participants with two scenarios about inclusion of surveys conducted as part of an outbreak investigation, resulting in a total of eight different scenarios about the interpretation of serological data. Specifically, those studies were included in scenarios 1-4 and excluded in scenarios 5-8.

#### Regression models

There are numerous methods for regression modeling, none of which is guaranteed to be optimal in any given application. To explore a range of regression models, we considered a total of eight that differed in terms of functional relationships between predictor and response variables and whether they allow for explicit spatial dependence. The first was the simplest possible model, in which we estimated only a single parameter describing a constant force of infection (FOI) across administrative units. Regardless of whether this model would perform well or not, we viewed it as a necessary benchmark against which other models should be compared. The second and third were linear regression models, one with only linear predictors and another with linear predictors and all possible two-way interactions thereof. The fourth and fifth were Markov random field models with no predictor variables but different numbers of free parameters (100 or 400) controlling the granularity of the spatial surface (10×10 or 20×20) that these models estimate. The sixth was a Markov random field model with 100 free parameters plus linear effects of predictors. All Markov random field models were implemented using the mgcv package in R [67]. The seventh was a random forest model, implemented in R with the randomForest package [68]. The eighth was a boosted regression trees model, implemented in R with the gbm package [69]. We included the full set of predictors in every model that made use of predictors, and we did not perform model selection to reduce the number of predictors. We made this choice given that our approach was already very computationally intensive and our motivating interest was in comparison of models that differ in structure rather than variable composition.

## Data Availability

Data are available upon request.

## ACKNOWLEDGEMENTS

We thank Tini Garske for sharing the outbreak dataset, Arran Hamlet for sharing estimates of population and vaccination coverage, and Freya Shearer and Simon Hay for sharing a raster for non-human primate occurrence probability.

## SUPPLEMENTAL APPENDIX

### Probabilities of different infection outcomes

There were two steps in our analysis for which we needed to make assumptions about the probabilities of different infection outcomes. In Step 2, we placed priors on the proportion of reported events that were cases or deaths. In the final step of translating ensemble projections of force of infection into deaths, we made an assumption about the proportion of infections that result in death.

We based both of these assumptions on a re-analysis of data compiled by Johansson et al. [3]. While we were amenable to using estimates of the probabilities of different infection outcomes by Johansson et al., information was only presented in that paper about marginal distributions of the probability of each infection outcome. We felt that it was important to make use of estimates for which correlation structure among the probabilities of different infection outcomes was accounted for.

A natural distribution for representing uncertainty in multiple probabilities that together sum to one is a Dirichlet distribution [70]. Accordingly, we used maximum likelihood to estimate parameters of a Dirichlet distribution describing the probabilities of the same four infection outcomes considered by Johansson et al.: A = asymptomatic infection; M = mild symptomatic infection; S = severe symptomatic infection; and F = infection resulting in a fatality. We used all data presented in Tables 1 and 2 of Johansson et al. in which two or more combinations of infection outcomes were reported in the same row. In some cases, this included sums of infection outcomes; namely, A+M and M+S, given ambiguity in some studies about these infection outcomes. To do this, we leveraged the property that the four concentration parameters of the Dirichlet distribution— α_*A*_, α_*M*_, α_*S*_, and α_*F*_ —can be summed to obtain concentration parameters of a lower-dimensional Dirichlet distribution (including a beta distribution in the case of two outcomes) that are consistent with the higher-dimensional Dirichlet distribution [70].

Our analysis resulted in maximum-likelihood estimates (MLE) of α_*A*_ = 8.69, α_*M*_ = 2.74, α_*S*_ = 4.10, and α_*F*_ = 2.05. This corresponds to mean estimates (and 95% credible intervals, by which we mean the 0.025-0.975 quantile range under the MLE Dirichlet parameters) of the probability that an infection results in outcomes of A, M, and S+F of 0.49 (0.27-0.72), 0.16 (0.03-0.35), and 0.35 (0.15-0.58), respectively. This compares with estimates by Johansson et al.[3]0.55 (0.37-0.74), 0.33 (0.13-0.52), and 0.12 (0.05-0.26), respectively. For the probability of death given severe disease, we obtained a mean estimate (and 95% credible interval) of 0.33 (0.04-0.57), as compared to an estimate of 0.47 (0.31-0.62) by Johansson et al. Together, this meant that our mean estimate of the probability of death upon infection of 0.12 was twice that of Johansson et al. (0.05), although there was also wider uncertainty in our estimate (0.01-0.27) than that of Johansson et al. (0.02-0.12). The differences between our estimates could be due either to the different distributions we used (Dirichlet vs. binomials) or the additional step in the analysis by Johansson et al. that estimated the prevalence of infection in the outbreaks from which the underlying data came.

### Variance partitioning

Given our interest in evaluating the sensitivity of disease burden and vaccination impact to model assumptions, quantifying the proportion of variance in those measures attributable to different sources was an important focus of our results. To do that, we applied the law of total variance [71] to draws of outputs of interest—force of infection (FOI), deaths averted, and deaths—from their posterior distributions.

For a given output of interest (e.g., force of infection), *Y*, and a single explanatory factor (e.g., first-level administrative unit, adm1), *X*, total variance in *Y* can be partitioned according to

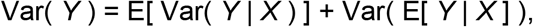

where the first term on the right-hand side represents variance in *Y* not accounted for by *X* (which we referred to as statistical uncertainty) and the second term represents variance in *Y* that is accounted for by *X*. This equation was used to calculate the proportion of variance in log_10_ FOI (*Y*) attributable to adm1 (*X*) in Step 3 and regression model (*X*) in Step 4.

In the case of two explanatory factors, *X*_1_ and *X*_2_, the above equation can be extended to

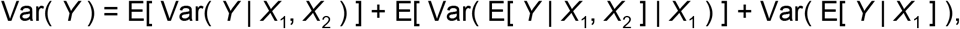

where the first term on the right-hand side represents variance in *Y* not accounted for by *X*_1_ or *X*_2_, the third term represents variance in *Y* accounted for by *X*_1_, and the second represents variance in *Y* accounted for by *X*_2_ conditional on *X*_1_ [72]. This equation was used to calculate the proportion of variance in log10 FOI (*Y*), deaths averted (*Y*), and deaths (*Y*) attributable to adm1 (*X*_1_) and serology scenario (*X*_2_). We selected adm1 as *X*_1_ given that spatial heterogeneity in yellow fever due to heterogeneity in underlying environmental drivers is considerable, making it logical to consider it as a primary source of variation in yellow fever’s burden.

## SUPPLEMENTAL FIGURES AND TABLE

**Figure S1.**
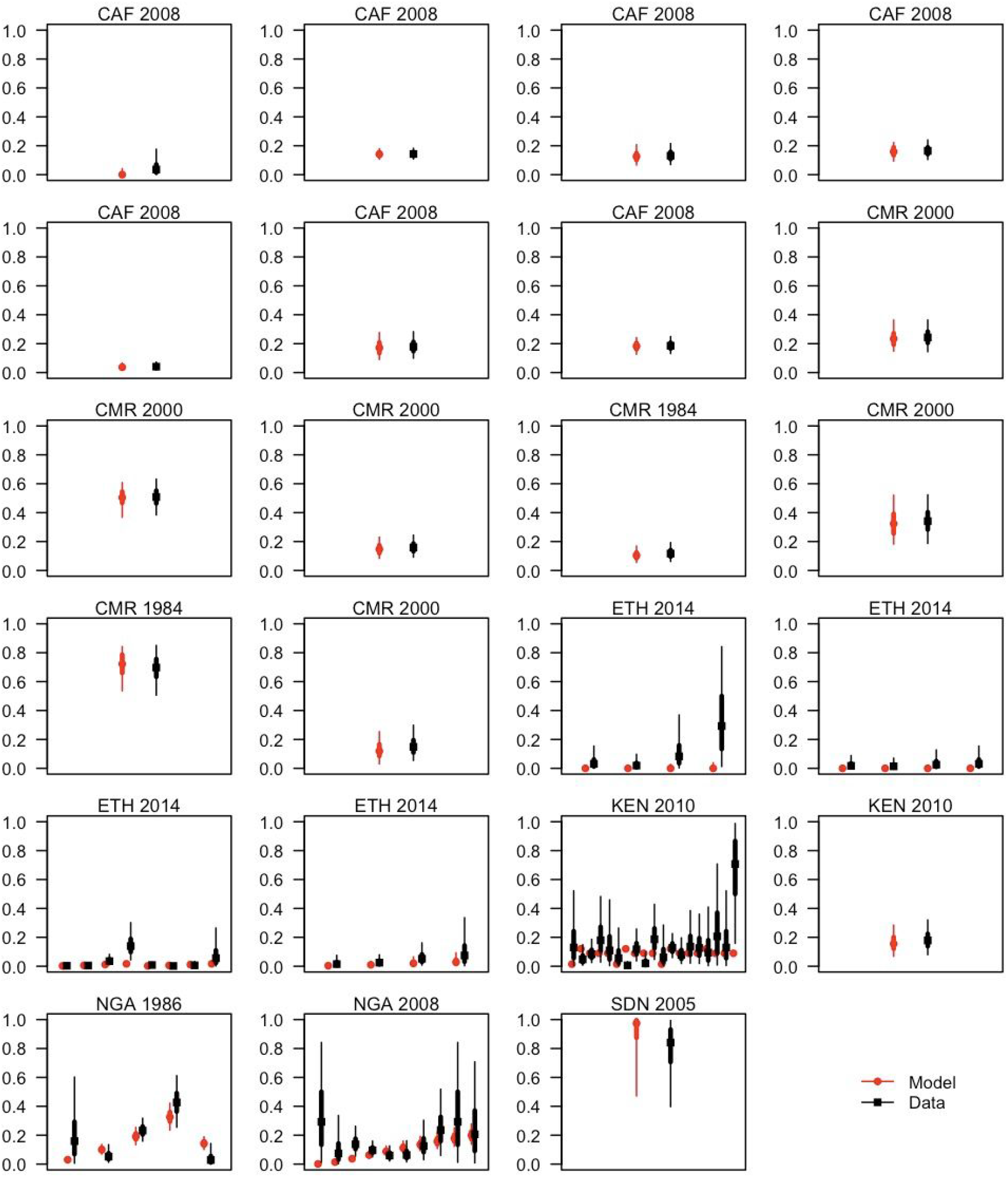
Comparison of seroprevalence based on estimates of force of infection from Step 1 (red) and simple estimates of seropositivity based on conjugate prior relationships (black) under serology scenario 1. The conjugate prior estimates assumed a flat beta prior with shape parameters equal to 1 and a binomial likelihood applied independently to each data point [66]. Within each panel, different pairs of estimates correspond to different age strata. Points indicate median, thick line segments indicate 50% posterior predictive intervals (PPIs), and thin lines indicate 95% PPIs. See Table S1 for more information about these studies. Results for other serology scenarios were similar and are not shown.

**Figure S2.**
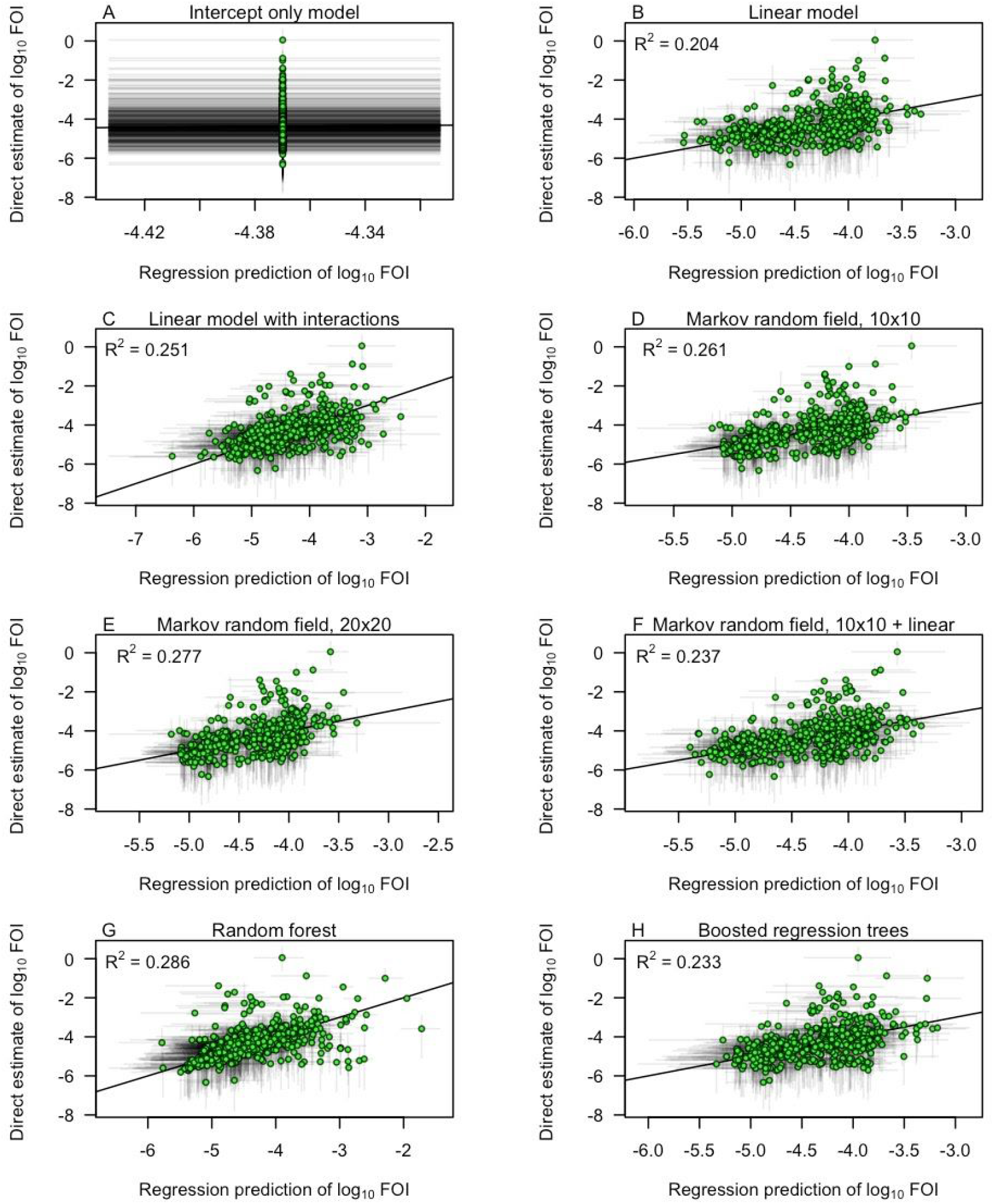
Comparison of regression predictions of force of infection from Step 4 (x-axis) against projected values from Step 3 (y-axis). Green circles indicate median values, and gray line segments indicate 95% uncertainty intervals. The coefficients of determination, R^2^, in each panel were calculated based on median values.

**Figure S3.**
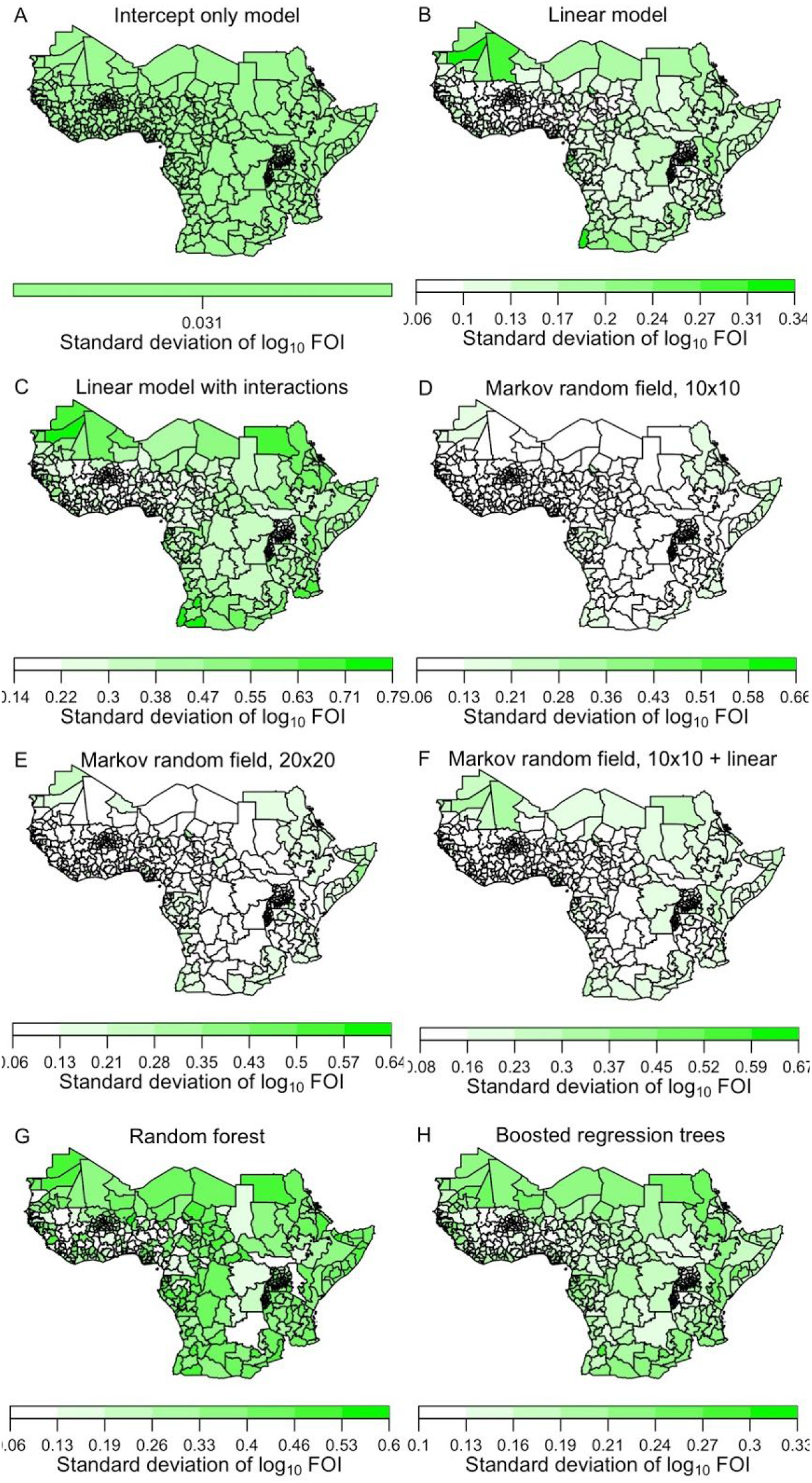
Uncertainty in spatial prediction of force of infection from eight regression models. Standard deviations of values on a log_10_ scale are shown from serology scenario 1. Color axes for each model differ so as to maximize contrast within each panel. Other serology scenarios produced similar results, but with magnitude varying according to differences in the estimated average reporting probabilities shown in Fig. 3.

**Figure S4.**
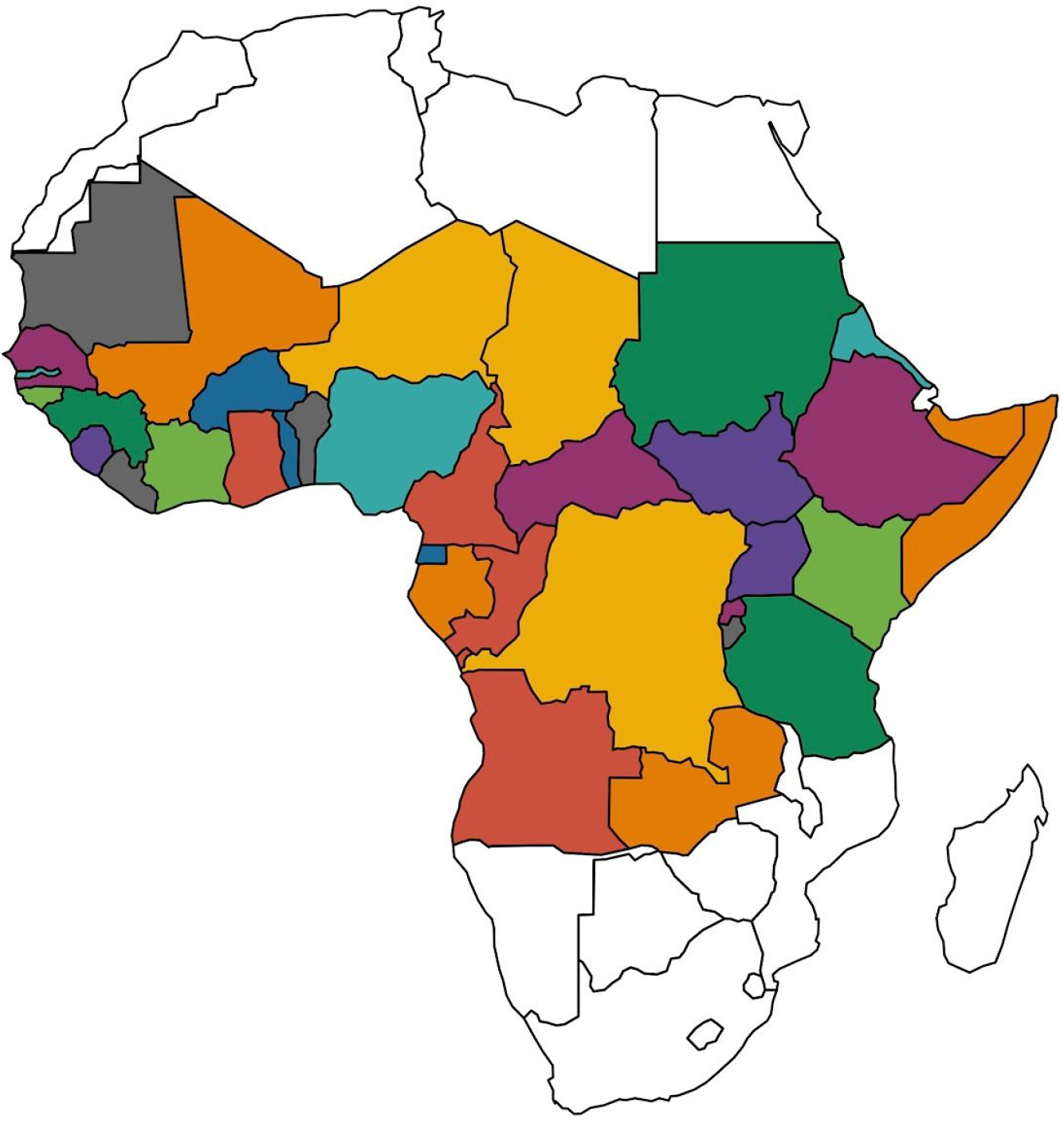
Country partitioning for cross-validation. Each color signifies a different set of countries for which data was withheld from model fitting and used to assess out-of-fit prediction. This partitioning was determined so as to maximize the evenness of the number of first-level administrative units (adm1s) across partitions. To do this, the first partition took the country with the most adm1s and the two countries with the fewest and grouped them together (i.e., Uganda, South Sudan, and Sierra Leone in purple). The second partition took the country with the second most adm1s and the two countries with the third and fourth fewest adm1s (i.e., Burkina Faso, Togo, and Equatorial Guinea in dark blue). This process was repeated until ten partitions of three to four countries each were obtained. Countries in white were not included in our analysis.

**Figure S5.**
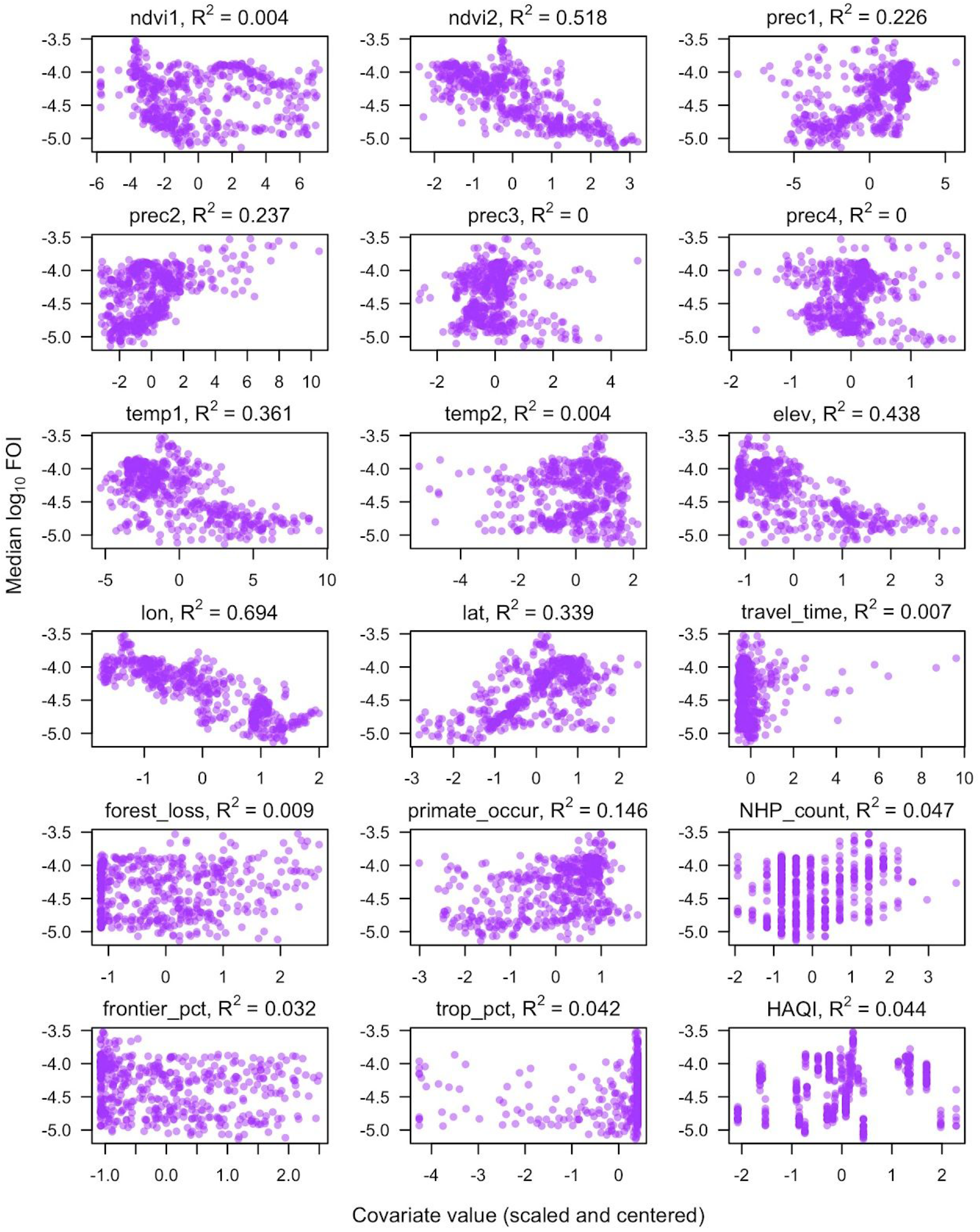
Associations between spatial covariates and log_10_ force of infection. Each dot corresponds to an adm1, with median values shown here and used to compute coefficients of determination, R^2^. All spatial covariates were centered and scaled for this and other analyses.

**Figure S6.**
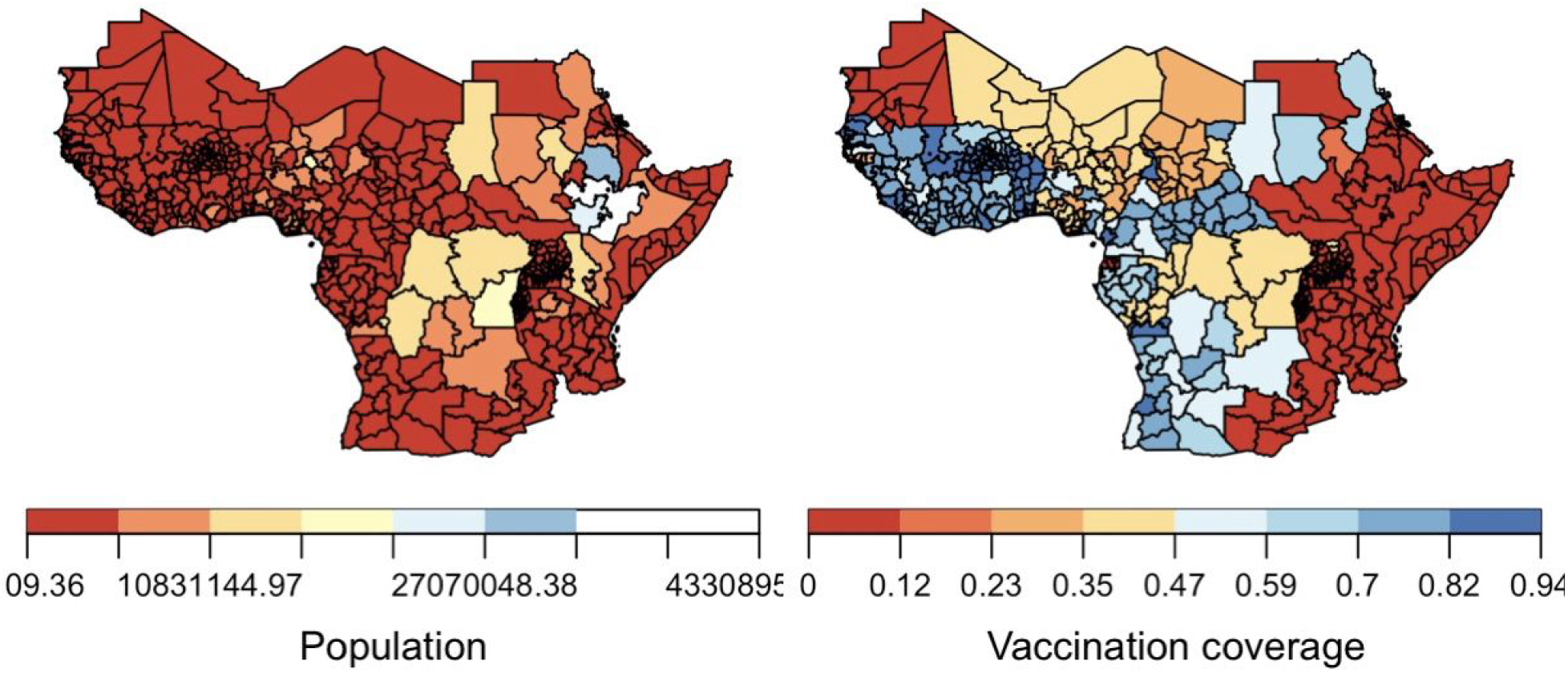
Population and vaccination coverage maps. Both maps reflect an average across 2021-2030. In the case of vaccination coverage, age-specific values of vaccination coverage were averaged proportion to population by age. All population and vaccination coverage estimates used in this analysis were generated by Hamlet et al. [8].

**Figure S7.**
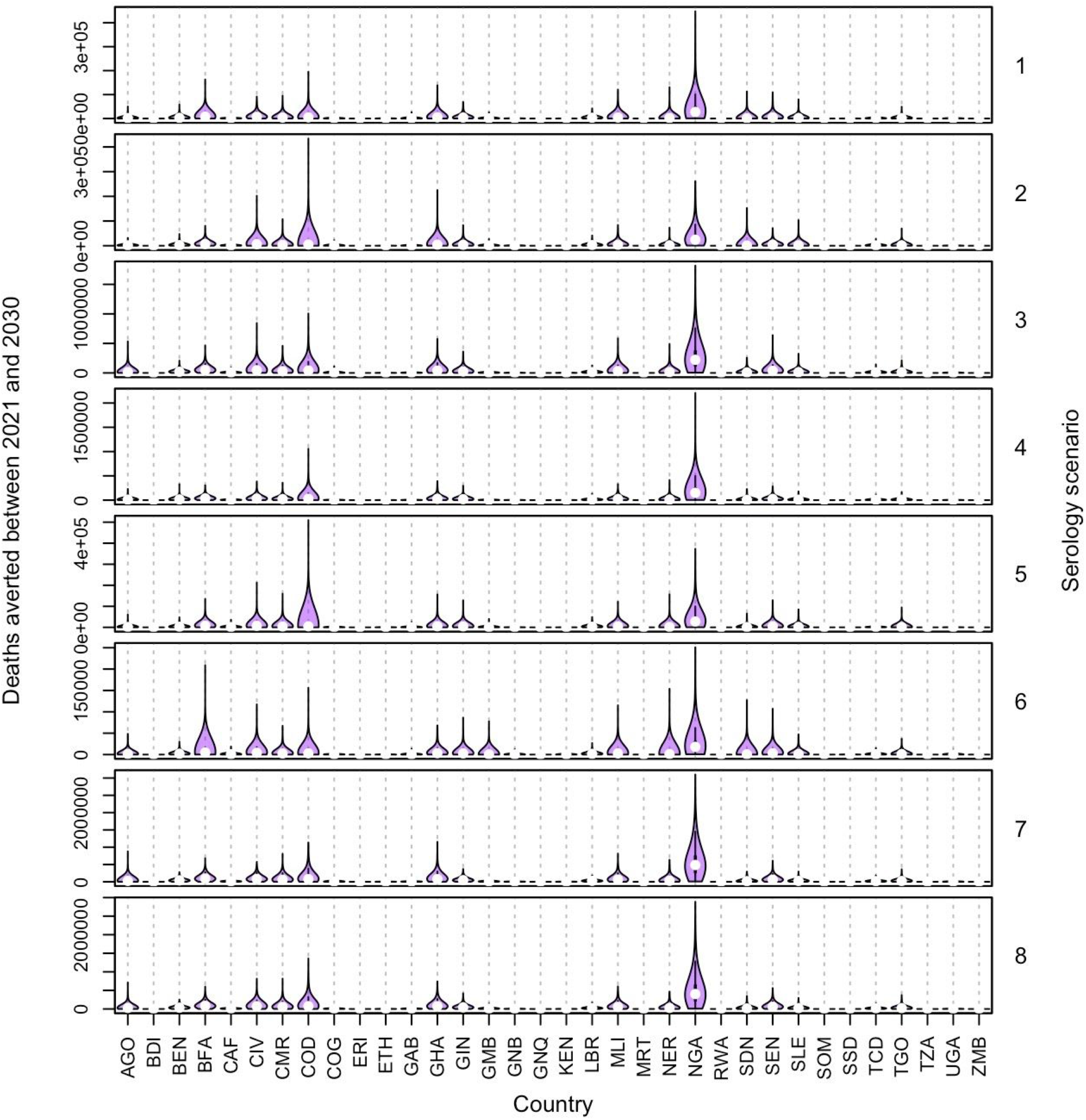
Deaths averted by country during 2021-2030. Results from the ensemble model corresponding to each serology scenario are shown in each panel.

**Figure S8.**
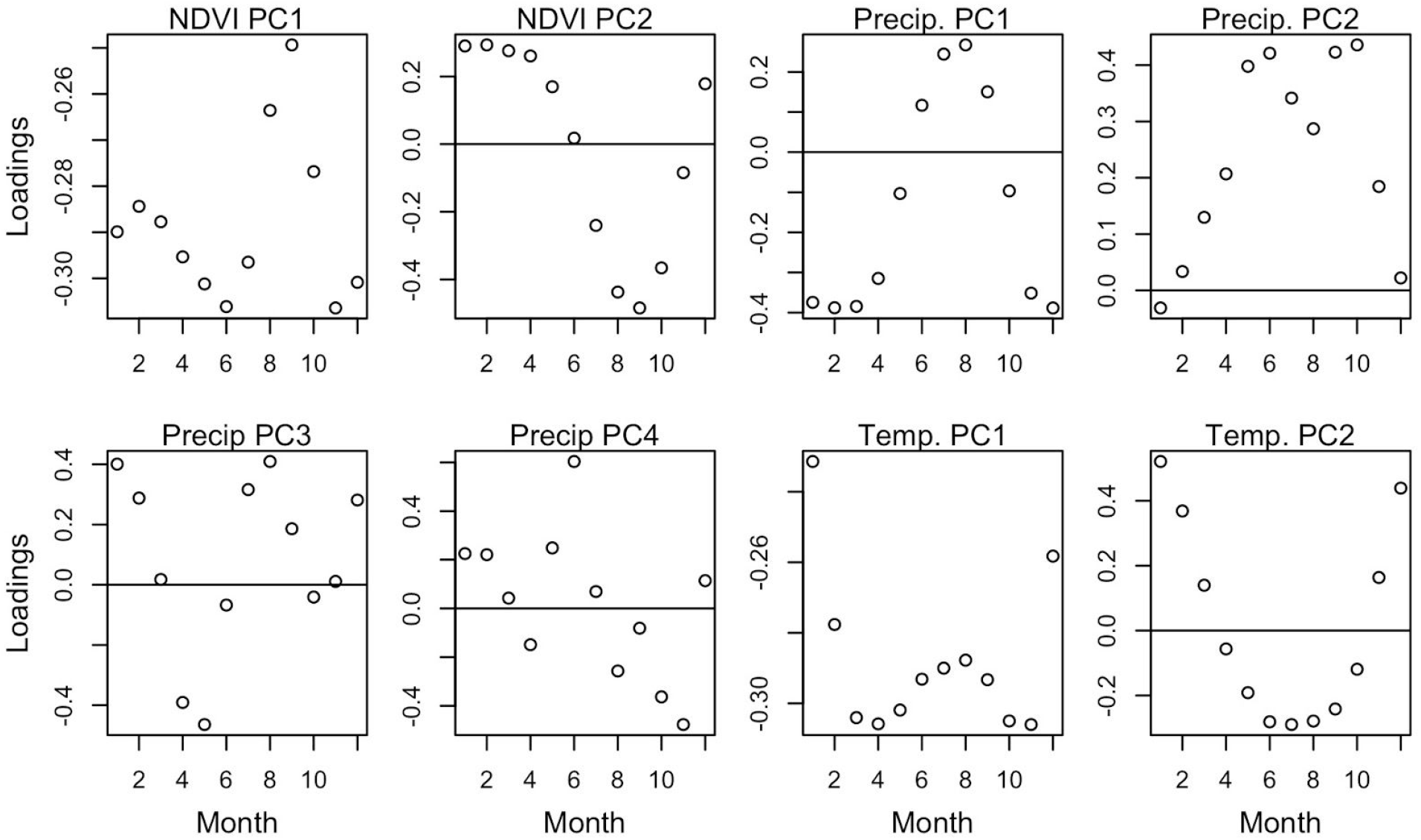
Principal component loadings for spatial covariates with monthly values. Each principal component results from summing the product of these loadings and the monthly values of a given spatial covariate for each administrative unit. For example, NDVI PC1 results from taking the loadings in the upper left panel, multiplying them by monthly NDVI values for each administrative unit, and then taking the corresponding sums. This reduces the dimensionality of the twelve monthly values of these three variables (a total of 36 variables) down to the eight represented here. The number of principal components for each of NDVI, precipitation, and temperature was chosen such that 95% of variation in those variables was accounted for by these principal components. Maps of the resulting principal components are displayed in Fig. S9. As can be gleaned from the loadings, many of these principal components appear to capture differences in seasonal climatic patterns across the study region.

**Figure S9.**
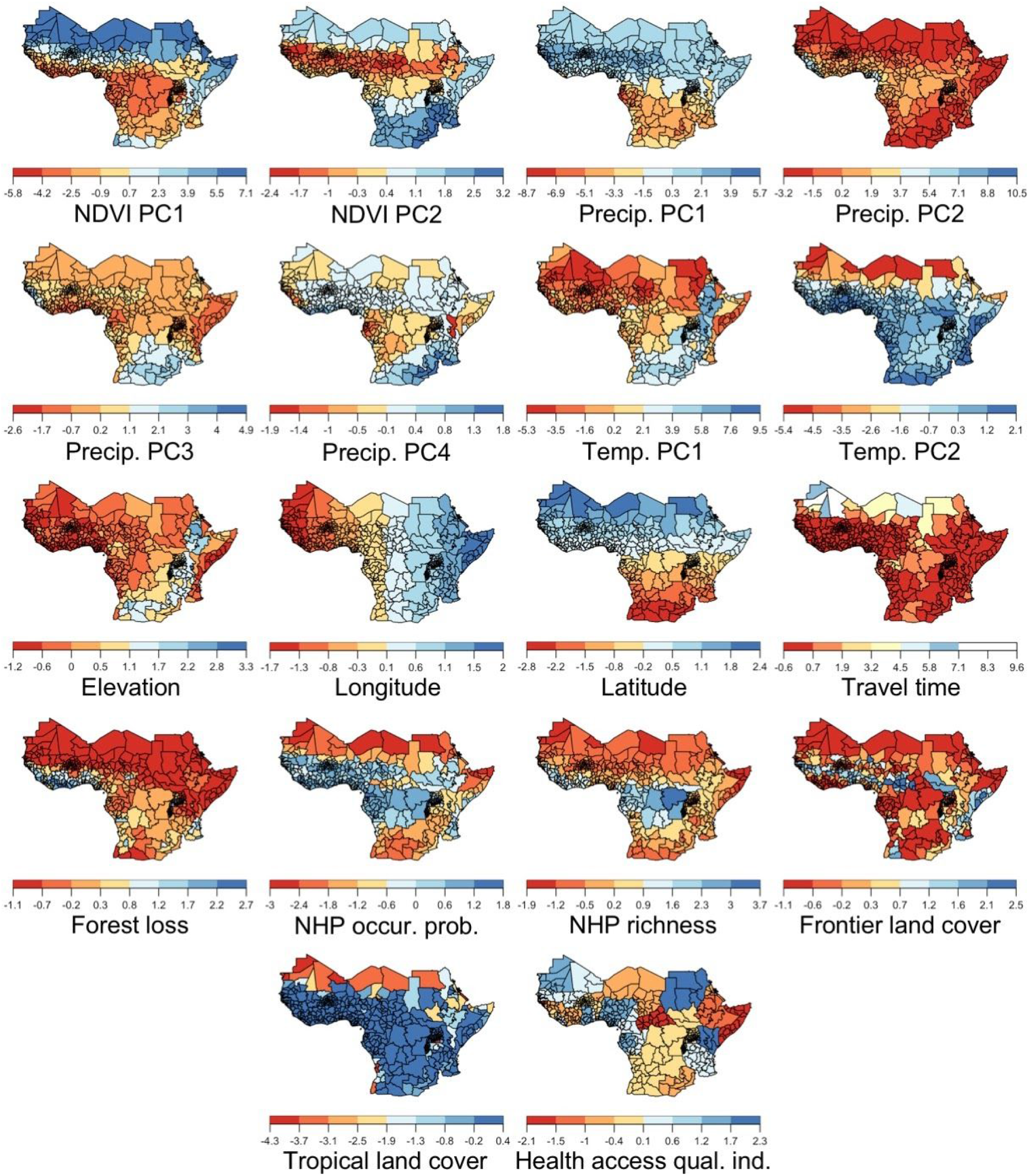
Spatial covariates used in the regression analysis in Step 4. All variables shown here have been centered and scaled, which is how they were used in the regression analysis and is sufficient to convey their relative spatial patterns. Rather than making inferences about relationships between these variables and force of infection, our goal was ensuring that we had variables with sufficiently diverse spatial patterns that the regression models would have sufficient flexibility to capture patterns in force of infection projected from Step 3.

**Table S1.** Characteristics of serological studies. Columns include characteristics that: allow for cross-referencing with Fig. 2, Fig. S1, and Fig. 3 (country, year, order therein); are related to how the serology scenarios were defined (vaccination status, vaccination coverage, outbreak investigation); determine force of infection (seropositive, number tested); and determine estimates of site-specific reporting probabilities (reported cases, deaths). Note that the number seropositive and number tested are totals across all age groups, which varied across studies and are not shown in this table in the interest of space. Vaccination coverage pertains to the year of the study and reflects a population-weighted average across age groups in the study.

| Country | Year | Vacc. status | Vacc. cov. | Outbreak investig. | Sero. pos. | Sero. tested | Cases | Deaths | Ref. |
| --- | --- | --- | --- | --- | --- | --- | --- | --- | --- |
| CAF | 2008 | No | 0.37 | No | 0 | 18 | 0 | 0 | [54] |
| CAF | 2008 | No | 0.39 | No | 48 | 339 | 0 | 0 | [54] |
| CAF | 2008 | No | 0.40 | No | 10 | 80 | 0 | 0 | [54] |
| CAF | 2008 | No | 0.39 | No | 18 | 112 | 0 | 0 | [54] |
| CAF | 2008 | No | 0.39 | No | 8 | 211 | 0 | 0 | [54] |
| CAF | 2008 | No | 0.44 | No | 11 | 64 | 2 | 0 | [54] |
| CAF | 2008 | No | 0.37 | No | 30 | 164 | 0 | 0 | [54] |
| CMR | 1984 | No | 0.20 | Yes | 10 | 90 | 3 | 2 | [55] |
| CMR | 1984 | No | 0.36 | Yes | 17 | 24 | 8 | 2 | [55] |
| CMR | 2000 | No | 0.28 | No | 13 | 55 | 4 | 0 | [56] |
| CMR | 2000 | No | 0.19 | No | 30 | 59 | 3 | 0 | [56] |
| CMR | 2000 | No | 0.19 | No | 13 | 85 | 0 | 0 | [56] |
| CMR | 2000 | No | 0.27 | No | 9 | 27 | 4 | 1 | [56] |
| CMR | 2000 | No | 0.19 | No | 4 | 30 | 2 | 0 | [56] |
| ETH | 2014 | No | 0.00 | No | 0 | 64 | 6 | 0 | [57] |
| ETH | 2014 | No | 0.00 | No | 0 | 135 | 0 | 0 | [57] |
| ETH | 2014 | No | 0.00 | No | 8 | 1,313 | 0 | 0 | [57] |
| ETH | 2014 | No | 0.00 | No | 2 | 152 | 0 | 0 | [57] |
| KEN | 2010 | Unknown | 0.06 | No | 22 | 433 | 119 | 46 | [58] |
| KEN | 2010 | Unknown | 0.01 | No | 6 | 36 | 0 | 0 | [58] |
| NGA | 1986 | No | 0.36 | Yes | 37 | 207 | 1,295 | 564 | [59] |
| NGA | 2008 | No | 0.29 | No | 25 | 310 | 407 | 46 | [60] |
| SDN | 2005 | No | 0.06 | Yes | 3 | 3 | 0 | 0 | [61] |

